# Reducing mental health inequalities through hypothetical interventions in early childhood: Evidence from mediation analysis using the UK Millennium Cohort Study

**DOI:** 10.64898/2026.01.07.26343647

**Authors:** Yu Wei Chua, Philip McHale, Daniela Schlüter, Viviane S. Straatmann, Eric T.C. Lai, Melisa Campbell, Bethan Plant, Michelle Black, David Taylor-Robinson

## Abstract

The early childhood environment influences later mental health, but it remains unclear which aspects are most important for reducing inequalities. We quantified how socioeconomic inequalities in mental health at 5 and 17 years (y) would change under hypothetical interventions on five aspects of the early childhood environment. We analysed parent-reported data from the UK Millennium Cohort Study (n=15513). Socioeconomic circumstances (SEC) were assessed using maternal education at 9 months. Mental health was measured using the Strengths and Difficulties Questionnaire total difficulties score at both ages. Five mediator pathways – neighbourhood, social support, material home environment, household adversity, and parent-child relations – were identified through stakeholder consultation and literature. Interventional Disparity Measures (IDMs) estimated how mental health inequalities would change if the confounder-adjusted mediator distributions for children in low SEC were shifted to match those in high SEC, using Monte Carlo simulations with imputed missing data. Mental health difficulties were higher among children in the low SEC (Primary or Lower) compared to high SEC (Tertiary) group (5y: 3.3[95%CI: 3.0;3.6] points; 17y: 3.1[2.8;3.5] points). Shifting all five pathways reduced these inequalities by around one-third (5y: −32%[−40;−23]; 17y: −35%[−48;−26]). At 5y, most of the inequality reduction was attributable to shifts in parent-child relations (−22% [−27;−16]). At 17y, both parent-child relations (−15%[−22;−9]) and social support (−11%[−21;−6]) were equally important. To reduce socioeconomic inequalities in mental health, policymakers should prioritise early interventions that improve parent-child relations and access to social support, while recognising the broader influence of neighbourhood, material, and household psychosocial contexts on these pathways.

---

Children and adolescents in families with low socioeconomic status are two to three times more likely to develop mental health problems than their more advantaged peers (1). Socioeconomic inequalities in children’s mental health emerge early and are well established before the age of 5 years, with little change over childhood and adolescence (2,3). This underscores the need to understand the factors in early childhood driving these inequalities, to target preventative interventions.

Previous research suggests that early childhood factors may account for up to two-thirds of socioeconomic inequalities in mental health. Studies on individual factors highlight the mediating role of the neighbourhood environment (4), parental mental health (5,6), adverse childhood experiences (7,8), childcare uptake (9,10), home learning environment (11), and parenting behaviours (12). Although these findings identify important domains, we lack an integrated understanding of the causal pathways through which inequalities arise. Organising early years determinants by level (child, parent, household, services, community), type (material, psychosocial, behavioural, educational), and timing (perinatal vs early childhood) may help policymakers navigate this evidence. Conceptual frameworks also emphasise that distal social determinants act through increasingly proximal factors and that inequalities arise through the unequal distribution and consequences of risk (13–15).

Few observational studies have compared multiple pathways simultaneously. Two key studies highlight the dominant role of psychosocial factors, particularly parental mental health, with smaller contributions from material and educational environments (16,17). However, these studies did not include key adversities such as exposure to domestic violence, known to explain a substantial proportion of socioeconomic inequalities in mental health (7,8). Previous studies have also assessed different pathways alongside the early childhood environment, to achieve broader research aims on understanding early life risk factors. However, this can influence conclusions on the importance of early childhood environmental pathways. For example, Straatmann and colleagues (2019) identified that the perinatal pathway was most important for inequalities in child mental health, yet several early-childhood pathways combined explained a similar proportion of inequalities (17). Additionally, a comparison with child sociodemographic characteristics - factors that are less modifiable - or measures of child developmental abilities (e.g., cognition) which are partly shaped by the environment, may underestimate the role of environmental pathways.

In addition to the limits of our understanding of the importance of different causal pathways, there are also gaps in the evidence on interventions to reduce inequalities in children’s mental health. Although programmes in early education, parenting, social protection, and (in low-income settings) nutrition and sanitation have shown effectiveness (18–23), evaluations typically focus on single interventions and use heterogeneous methods, making synthesis difficult (18). The need for longitudinal follow-up also means impacts on adolescent mental health take years to assess (24). A further gap is that evaluations are limited to examining inequality impacts of specific interventions. However, policymakers also need stronger evidence on the critical ages and the specific nature of interventions needed to reduce inequalities in mental health at scale. This is key to prioritise resources towards the most effective time to intervene, and understand the most effective interventions, which may vary at different developmental periods as children transit into different settings and environments.

Observational studies offer a way to simulate the impact of potential interventions that have not yet been implemented and can provide rapid insights into real-world levers for reducing inequalities (24). Building on advances in organising early-years risk and protective factors (16,17), we use a nationally representative UK birth cohort study to examine the change in socioeconomic inequalities in mental health at 5 and 17 years if important aspects of the early childhood environment were hypothetically intervened on. We aimed to quantify the change in mental health inequalities by altering the distribution of early childhood environmental factors observed in the least advantaged to that of the most advantaged, using factors across five pathways (neighbourhood, social support, material home environment, household adversity and parent-child relational). We approached this in two steps, first quantifying the change in mental health inequalities when all pathways are shifted together; and second when each pathway is shifted individually.

## Methods

### Participants

The UK Millennium Cohort Study (MCS) is a longitudinal birth cohort study of children and their families born between 2000 and 2002, representative of the UK population (25). It includes survey questionnaires at 9 months (m), 3, 5, 11, 14, 17 and 23 years (y) to date. Ethical approval for the MCS was provided by an NHS Research Ethics Committee, in accordance with the 1964 Declaration of Helsinki and its later amendments or comparable ethical standards (25). No further ethical approval was required.

Participants with complete data on the exposure and at least one parent or self-reported Strengths and Difficulties Questionnaire (SDQ) measure from 5y to 17y were included in the main analytic sample (n=15513), with any missing mediators, covariates and outcomes to be imputed (Figure 1). The 5y (n=8477) and 17y (n=5612) complete case samples included participants with complete data on the exposure (9m maternal education), mediators, covariates and outcome data at 5y or 17y, respectively.

**Figure 1.**
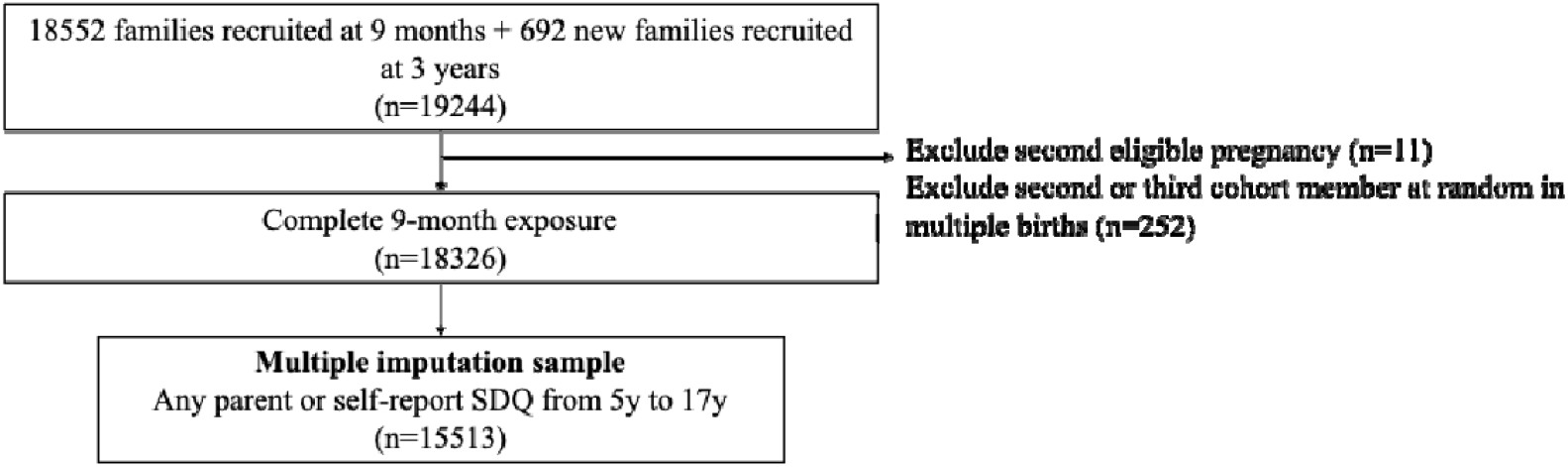
STROBE flowchart showing inclusion and exclusion criteria for defining the sample for the present analysis.

### Patient and public involvement

To bridge academic understanding with practice, we involved two public health professionals working in commissioning of 0 to 19 years services in Liverpool and Sheffield City Councils in a research consultation on 27 March 2025 and in drafting the manuscript. The research consultation explored the relevance of the research questions, appropriateness of variables and characterisation of the early childhood environment.

### Outcomes

Our primary mental health outcomes at 5y and 17y were parent report of the total difficulties score on the SDQ (26). The SDQ is a validated questionnaire comprising five factors: hyperactivity, conduct, emotional, peer problems and the prosocial scale. We used the total score (range: zero to 40), which is the sum of the four problem scales (range: zero to 20), operationalised as a continuous scale to capture information on mental health difficulties across all severity levels. Higher scores indicate more symptoms.

### Exposure

Our exposure was High, Middle or Low childhood socioeconomic circumstances (SECs) at 9 months, assessed using maternal highest academic qualification (High: Tertiary; Middle: Lower Secondary to Diploma/A level, and Low: Primary or lower). The categories were based on the International Standard Classification of Education (ISCED) 2011 (27,28) (Appendix S1)

### Mediators

Our selection of mediators was guided by prior research and participatory research (see earlier section on Patient and public Involvement). Mediators were initially grouped into pathways based on previous research on the level and type of intervention, and distal-to-proximal ordering (7,8,12,13,16,17). Following feedback from public health professionals, we modified the grouping, naming, order, and variables characterising the paths (See Appendix S1). Therefore, this approach integrates previous understanding on key pathways in the early childhood environment with stakeholder perspectives, to provide an organisational framework that supports the translation of evidence into practice.

The resulting five mediating early childhood pathways (a–e) assessed at 9m or 3y were: the neighbourhood, social support, material home environment, household adversity and parent-child relational pathways (Box 1, with operationalisation of each variable detailed in Appendix S2). All mediator variables were caregiver reported apart from two interviewer-assessed variables of the material home environment.

#### Box 1. Early childhood environment mediating pathways

**Early childhood environment pathways**

a. **Neighbourhood** This pathway was defined as the wider physical environment and structural factors that impact an individual’s opportunity. Five binary variables were reduced to a two-class variable using latent-class analysis (see Appendix S1, Table S1 and Figure S1),
  1. Neighbourhood environment (presence of environmental problems, rubbish lying around)
  2. Neighbourhood accessibility (presence of poor public transport, poor access to shops)
  3. Neighbourhood crime (presence of racism, vandalism)
  4. Perception of living in a safe neighbourhood
  5. Availability of safe places for children to play
  Two class variable obtained from latent class analysis: Good / Poor neighbourhood conditions.
b. **Social support** This pathway was defined as the wider social environment, including access to early interventional services and social support from friends or families. Proxied using three binary variables:
  1. Attendance of antenatal classes (to proxy early engagement with healthcare services)
  2. Early access of centre-based childcare
  3. Low social support
c. **Material home environment** This pathway was defined as the physical aspect of the home environment, including housing conditions, play environment (such as books, toys, space), and other material goods that shape children’s growth. Seven binary variables were used to capture this pathway, reduced to a three-class variable using latent class analysis (see Appendix S1, Table S1 and Figure S1).
  1. Not able to afford essentials for child
  2. Problems with heating
  3. Problems with damp
  4. Household overcrowding
  5. Insecure housing
  6. Safe in-home play environment (interviewer assessed)
  7. Parent provided toys during visit (interviewer assessed)
  Three-class variable was characterised by Good housing and no overcrowding / Overcrowding with lower risk of poor housing / Poor housing with lower risk of overcrowding. Overcrowding was also characterised by low availability of toys at the visit.
d. **Household adversity** This pathway was defined as the psychosocial issues within the wider household unit characterised by poor parental mental health or dysfunctional household social interactions. This can be considered a more distal emotional pathway together with the parent-child relational pathway.
  1. Main or partner respondent experience of domestic violence (binary)
  2. Main or partner respondent substance misuse (binary)
  3. Main respondent mental distress (Continuous, higher score indicates greater distress)
e. **Parent-child relational** This pathway was defined as the parenting behaviours that influence parent-child interactions and relationships. This can be considered a more proximal emotional pathway together with the household adversity pathway, and the most proximal pathway as the early caregiver-child psychosocial interactions play a major role in shaping children’s development in early life.
  1. Frequency of parenting activities (Continuous, higher score indicates greater frequency)
  2. Disciplinary parenting (Continuous, higher score indicates greater use of disciplinary practices)
  3. Parent-child relationship (Continuous, Higher score indicates more positive parent-child relationship)

### Covariates

All covariates were assessed using parent report at 9m, except longstanding child illness (assessed at 3y) and parental alcohol misuse (assessed at 9m or 3y; Appendix S3).

Baseline confounders of the exposure–outcome (E–O) relationship included parental demographic and life-history factors occurring before the child’s birth—ethnicity, parental mental health history, parental childhood adversity (proxied by being in care), and maternal age—as these influence both socioeconomic circumstances and child mental health.

Confounders of each mediator–outcome (M–O) relationship were identified separately for each pathway. For each M–O model, we adjusted for its specific M–O confounders, all baseline confounders, and any variables in preceding pathways. We included cultural differences (language spoken at home) and number of children in the household - as these shape caregiver stress, material resources, parental time, and parenting behaviour. We also adjusted for post-exposure intermediate confounders related to early child characteristics (gestational age, early developmental delays, longstanding limiting illness), which may affect exposure or vulnerability to early-childhood mediators via impacts on costs, parenting stress, and early parent–child activities. Parental alcohol misuse - excluded from the mediator set due to prior evidence of a reverse social gradient in the MCS (8) - was controlled for as an intermediate confounder. Child sex and other individual characteristics were included as effect modifiers of E–O and M–O relationships.

Auxiliary variables at 9 months or 3 years (voting status, breastfeeding history, accommodation type, housing tenure, occupational status, family structure, and household income) were included in the imputation models as they are variables which predict missingness in later waves (29).

### Framework and logic model

The hypothesised relationships between mediating pathways, exposures, outcomes and confounding relationships are shown in Figure 2. Mediator pathways were ordered from distal to proximal (14) for example, to capture the more plausible influence of the wider neighbourhood on social support available and the quality of the home and how the home environment could influence the severity of psychosocial issues and parenting activities.

**Figure 2.**
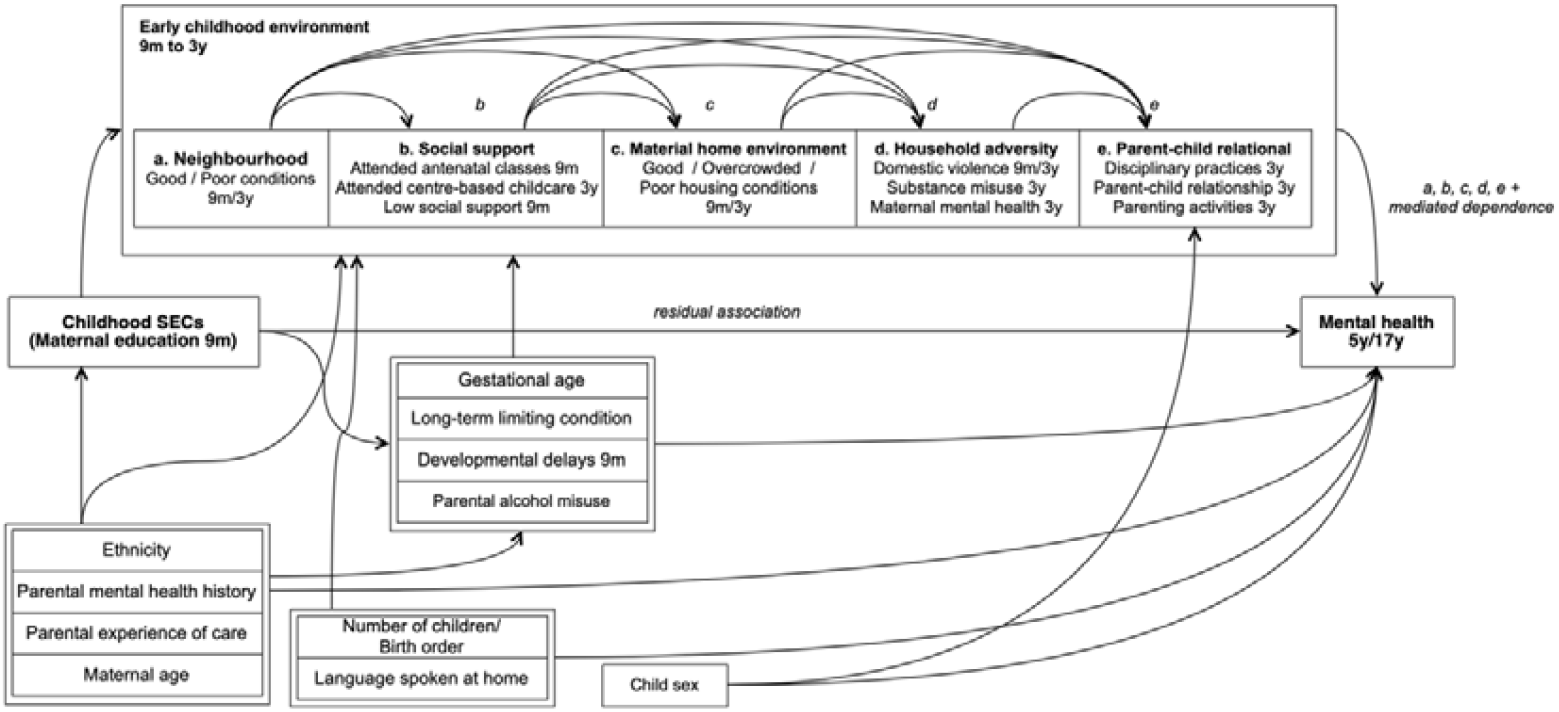
Directed Acyclic Graph showing hypothesised relationships between mediating pathways, exposures, outcomes and covariates. Child early developmental characteristics (Gestational age, long-term limiting condition, developmental delays 9m) were considered effect modifiers of the mediator-outcome relationship. Child sex was considered as an effect modifier of the E-O relationship, and for the M-O relationship for the Parent-child relational pathway.

Our conceptualisation of the early childhood environment recognises the complexity of early childhood pathways, which may influence subsequent mental health through various downstream pathways later. However, what we are interested in here is the difference in mental health outcomes if we were able to intervene on these early life mediators, given their connection with these complex downstream pathways which we do not further decompose (See Appendix S1).

### Interventional Disparity Measures (IDMs)

IDMs have been defined previously for multiple-mediator pathways (30,31). IDMs quantify changes in a health disparity due to a hypothetical mediator intervention, identified under the assumptions of 1) consistency, 2) no interference and 3) no unmeasured confounding of the mediator-outcome relationship. (See Appendix S4).

In this study, the inequality of interest is the difference in mental health outcome between Low and High SEC groups differing in maternal education, and the hypothetical intervention is a shift in the distribution of early childhood mediators (conditional on confounders) from what is observed in the Low SEC group to what is observed in the High SEC group. First, we simulate a scenario where mediators are intervened on *en bloc*, by randomly drawing each mediator from the joint distribution of all mediators. Secondly, we decomposed the pathways under scenarios where mediators in each pathway are individually shifted. IDM effects of interest are described in Box 2 and mathematical definitions provided in Appendix S5.

#### Box 2. Interventional disparity measures IDM effects of interest

- ***Adjusted Total Association (AdjTA)***. The confounder-adjusted total association between exposure and outcome. In this study, AdjTA refers to the difference in mental health outcome between Low versus High SEC groups, conditional on the confounders in Low and High SEC groups.
- ***Indirect Effect (IE)***. Change in the AdjTA after versus before the mediator intervention. IE here refers to the change in the difference in mental health outcome between Low vs High SEC groups when mediators are drawn from the High SEC distribution versus when mediators are drawn from the Low SEC distribution, conditional on the confounders in Low and High SEC groups.
- ***Residual Association (RA)***. Association between exposure and outcome remaining after the mediator intervention. RA here refers to the difference in mental health outcome that remains between Low vs High SEC groups, when both groups have mediators drawn from the High SEC distribution, conditional on the confounders in Low and High SEC groups. If additional assumptions are satisfied, this is equivalent to the Direct Effect in causal mediation.

**Decomposition of the IDM IE via five early childhood pathways**

- ***Mediator/pathway specific IE***. Change in the AdjTA due to a mediator intervention on that mediator/pathway, but not due to mediators/pathways downstream of it. Changes in the AdjTA acting as part of a chain of mediators are captured in the component of the most downstream mediator, as illustrated in Figure 2. (see Appendix S4 for further details on IDMs in the context of multiple mediators pathways)
- ***Mediated Dependence (MD)***. Change in the AdjTA due to a mediator intervention on one mediator pathway that modifies the effect of another pathway on the outcome. As this effect depends on the characteristics of two or more pathways, it is separately estimated as this effect cannot be attributed to one pathway alone.

### Statistical analysis

Statistical analyses were conducted in R (V4.3.2).

#### Descriptive statistics

We summarised missing data, described demographic characteristics and mediator distributions by socioeconomic circumstances for the main (imputation) analytic sample.

#### Exposure-outcome (E-O) and Mediator-outcome (M-O) associations

We estimated E–O and M–O associations using linear or logistic regression (*glm* function), adjusting for relevant confounders. To describe the main analytic dataset, we used multiple imputation (m = 10; *mice* package), including exposures, mediators, covariates, auxiliary variables, and the sampling stratum variable. Interaction terms were not included for interpretability. To describe complete case samples at 5 and 17 years, we fitted models using *svyglm* to incorporate design and non-response weights. Complete case samples have high missing data that non-response weights likely do not fully account for, as we required all exposure, mediators, outcome and covariates to be non-missing. Therefore, descriptive tables for complete case samples are reported in the Appendices.

#### Model fitting

Preliminary complete case analyses were used to fit outcome and mediator models required for estimating IDMs (Appendix S1). Regression assumptions were checked (Figures S2–S3). Due to skewed residuals, continuous mental health outcomes were square-root transformed; predictions were later back-transformed when estimating IDMs.

#### Monte Carlo simulation of IDMs with imputed data

We used the main analytic sample, imputing data once before the Monte Carlo simulation, before simulating outcomes based on hypothetical scenarios. We simulated hypothetical outcomes for individuals with High and Low SEC only, given mediator values drawn from the joint distribution of all mediators in the High or Low SEC distribution. To minimise Monte Carlo error, we simulated outcomes based on 200 draws of each mediator (Table S2), calculated IDMs based on the equations in Appendix S5, and marginalising over the resulting 3,102,600 Monte Carlo observations.

#### Decomposition via five pathways

For pathway-specific effects (indirect effect and mediated dependence term), mediator values were instead drawn from the distribution marginal to other pathways.

### Sensitivity analyses

To account for measurement differences in adolescent mental health, we repeated E–O and M–O models and IDM simulations using adolescent self-reported SDQ scores at 17y and parent-reported probable psychiatric disorder based on the SDQ threshold (Total Difficulties ≥17) (26). Additional methodological details are in Appendix S1. We also provide descriptive statistics of these outcome measures in the analytic sample.

## Results

### Missing data

In the main analytic sample (n=15513), 14192 (91.5%) of individuals had at least 2 SDQ measures and 6503 (41.9%) individuals had complete SDQ parent or self-reported data (Table S3). 1493 (9.6%) and 6640 (42.8%) individuals had missing data on parent-reported 5y and 17y parent-reported mental health respectively. Missing data on mediators ranged from 0% (neighbourhood and material latent class mediators) to 28.4% (disciplinary practices). Data on most confounders were complete, and some confounders with 2.4% to 11.1% missing data (Table S4).

### Sample characteristics

In the main analytic sample, prior to multiple imputationOfficial (Table 1), 2668 (17.2%), 10037 (64.7%), and 2808 (18.1%) fell in the High, Middle and Low SEC groups respectively. Parent-reported total difficulties score increased with greater socioeconomic disadvantage at 5y (mean(SD) High SEC: 5.4(4.0); Middle: 7.3(4.8); Low: mean(SD): 9.7 (5.7)) with similar levels at 17y. At 17y, a greater percentage of individuals met the SDQ threshold for a probable mental health disorder, increasing with socioeconomic disadvantage (n(%) High SEC: 70(3.6%); Middle: 516(9.1%); Low: 181(14.0%)). Self-reported 17y mental health difficulties were greater than parent-reported difficulties, with a narrower socioeconomic gradient (High SEC: 10.3[9.9;11.0]; Middle: 11.5[11.0;12.0]; Low: 12.9[12.0; 13.0]). Characteristics of the complete case 5y (n=8477) and 17y (n=5612) samples are provided in Table S5.

**Table 1.** Outcome and demographic characteristics by socioeconomic circumstances (analytic sample, before imputation)

| Characteristic | High, N = 2,668<sup>1</sup> | Middle, N = 10,037<sup>1</sup> | Low, N = 2,808<sup>1</sup> | p value |
| --- | --- | --- | --- | --- |
| 5y Parent-reported mental health ($n=14020$) | 5.4 (4.0) | 7.3 (4.8) | 9.7 (5.7) | <0.001 |
| Missing | 121 | 791 | 581 |  |
| 17y Parent-reported mental health ($n=8873$) | 5.6 (4.9) | 7.6 (5.9) | 9.5 (6.1) | <0.001 |
| Missing | 745 | 4,379 | 1,516 |  |
| 17y Self-reported mental health ($n=9315$) | 10.3 (5.1) | 11.5 (5.6) | 11.7 (5.8) | <0.001 |
| Missing | 656 | 4,148 | 1,394 |  |
| 17y mental health disorder positive screen (parent-report) ($n=8873$) | 70 (3.6%) | 516 (9.1%) | 181 (14.0%) | <0.001 |
| Missing | 745 | 4,379 | 1,516 |  |
| Household income (9 months) |  |  |  | <0.001 |
| 1 | 99 (3.7%) | 2,075 (20.7%) | 1,482 (53.0%) |  |
| 2 | 155 (5.8%) | 2,370 (23.7%) | 867 (31.0%) |  |
| 3 | 346 (13.0%) | 2,317 (23.1%) | 303 (10.8%) |  |
| 4 | 731 (27.4%) | 2,012 (20.1%) | 99 (3.5%) |  |
| 5 | 1,336 (50.1%) | 1,246 (12.4%) | 47 (1.7%) |  |
| Unknown | 1 | 17 | 10 |  |
| Income poverty (Below 60% median OECD equivalised household income) |  |  |  | <0.001 |
| Not in poverty | 2,501 (93.8%) | 6,772 (67.6%) | 782 (27.9%) |  |
| In poverty | 166 (6.2%) | 3,248 (32.4%) | 2,016 (72.1%) |  |
|  | 1 | 17 | 10 |  |
| Housing tenure |  |  |  | <0.001 |
| Owned | 2,380 (89.2%) | 6,347 (63.2%) | 915 (32.6%) |  |
| Rented/other | 288 (10.8%) | 3,688 (36.8%) | 1,893 (67.4%) |  |
| Unknown | 0 | 2 | 0 |  |
| Occupational status |  |  |  | <0.001 |
| Managerial/Professional | 2,155 (84.4%) | 3,036 (31.9%) | 182 (6.8%) |  |
| Intermediate | 109 (4.3%) | 1,246 (13.1%) | 96 (3.6%) |  |
| Small employers/self-employed | 90 (3.5%) | 739 (7.8%) | 184 (6.8%) |  |
| Lower supervisory/technical | 49 (1.9%) | 877 (9.2%) | 175 (6.5%) |  |
| Semi-routine/routine | 75 (2.9%) | 1,797 (18.9%) | 698 (25.9%) |  |
| Unemployed | 76 (3.0%) | 1,814 (19.1%) | 1,359 (50.4%) |  |
| Unknown | 114 | 528 | 114 |  |
| Family structure |  |  |  | <0.001 |
| Natural/reconstituted family | 2,529 (94.8%) | 8,254 (82.2%) | 1,933 (68.8%) |  |
| Lone parent | 139 (5.2%) | 1,783 (17.8%) | 875 (31.2%) |  |
| Sex |  |  |  | 0.130 |
| Female | 1,332 (49.9%) | 4,832 (48.1%) | 1,396 (49.7%) |  |
| Male | 1,336 (50.1%) | 5,205 (51.9%) | 1,412 (50.3%) |  |
| Ethnicity |  |  |  | <0.001 |
| White | 2,314 (92.7%) | 8,954 (93.0%) | 1,987 (76.9%) |  |
| Asian | 26 (1.0%) | 94 (1.0%) | 46 (1.8%) |  |
| Black | 95 (3.8%) | 183 (1.9%) | 88 (3.4%) |  |
| Mixed | 51 (2.0%) | 270 (2.8%) | 332 (12.9%) |  |
| Other | 11 (0.4%) | 129 (1.3%) | 130 (5.0%) |  |
| Unknown | 171 | 407 | 225 |  |
| Teenage mother at first live birth | 9 (0.3%) | 623 (6.2%) | 454 (16.2%) | <0.001 |
| Maternal age (years) | 31.9 (4.2) | 28.0 (5.9) | 26.8 (6.4) | <0.001 |
| Unknown | 0 | 0 | 2 |  |
| Maternal mental health history | 396 (14.8%) | 2,691 (26.8%) | 840 (29.9%) | <0.001 |
| Maternal experience of being in care | 9 (0.3%) | 126 (1.3%) | 112 (4.0%) | <0.001 |
| Children in household |  |  |  | <0.001 |
| 1 | 584 (23.4%) | 2,514 (27.8%) | 414 (17.9%) |  |
| 2 | 1,409 (56.4%) | 4,054 (44.8%) | 782 (33.8%) |  |
| 3 | 399 (16.0%) | 1,716 (19.0%) | 555 (24.0%) |  |
| 4 or more | 107 (4.3%) | 771 (8.5%) | 565 (24.4%) |  |
| Unknown | 169 | 982 | 492 |  |
| Gestational age (weeks) | 39.5 (1.9) | 39.4 (2.0) | 39.2 (2.1) | <0.001 |
| Unknown | 89 | 375 | 74 |  |
| Preterm birth (<37 weeks of gestation) |  |  |  | <0.001 |
| Preterm | 233 (9.0%) | 1,059 (10.9%) | 380 (13.5%) |  |
| Term | 2,363 (91.0%) | 8,672 (89.1%) | 2,428 (86.5%) |  |
| Unknown | 72 | 306 | 0 |  |
| Language spoken at home |  |  |  | <0.001 |
| English only | 2,260 (84.7%) | 9,073 (90.4%) | 2,011 (71.6%) |  |
| English and other languages | 408 (15.3%) | 963 (9.6%) | 797 (28.4%) |  |
| Unknown | 0 | 1 | 0 |  |
| Long-term limiting illness | 68 (2.7%) | 265 (2.9%) | 88 (3.8%) | 0.053 |
| Unknown | 187 | 1,030 | 510 |  |
| Developmental delays (9 month) | 391 (15.1%) | 1,462 (15.0%) | 490 (17.5%) | 0.006 |
| Unknown | 72 | 306 | 0 |  |
| Note:<br>N (%) and Pearson Chi-squared tests for categorical variables; Mean (SD) and Kruskal-Wallis rank sum test for continuous variables |  |  |  |  |

The distributions of mediators also show a gradient of greater proportion of risk factors and fewer protective factors with increasing socioeconomic disadvantage, except social support and disciplinary parenting (**Error! Reference source not found**.). A higher percentage of low social support was seen in the High SEC group (6.6%), compared to Middle (3.2%) and Low SEC (6.6%). High and Middle SEC groups show similar levels of disciplinary practices (mean(SD) High: 13.2(4.5); Middle: 13.1 (5.0)), while Low SEC groups show lower disciplinary practices (12.2 (5.5)). Distribution of mediators for the complete case samples can be found in Tables S6– S7.

### E-O and M-O associations

Controlled for baseline confounders, compared to High SEC, Individuals from Low SEC backgrounds had around 3 points greater parent-reported mental health difficulties at 5 and 17 years (Table 2). Controlled for the exposure, baseline, M-O confounders, and mediators in any preceding pathway, almost all mediators were associated with mental health difficulties. Good neighbourhood conditions, attendance of antenatal classes, attendance of centre-based childcare, positive parent-child relationships, and greater frequency of parenting activities were protective, while low social support, poor housing conditions or overcrowded conditions, presence of domestic violence, greater mental distress and substance misuse and greater disciplinary practices were risk factors (Table 2). Evidence for the positive association between substance misuse and parent-reported mental health difficulties was weak at both timepoints (5y: 0.22 points [−0.06; 0.51] p=0.126; 17y: 0.17 points [−0.26;0.6] p=0.421). Attendance of centre-based childcare was associated with lower mental health difficulties with a trend towards significance at 95% confidence level (5y: −0.23 points [−0.43; −0.05], p=0.057; 17y: −0.31 points [−0.63;0.02], p=0.066). Complete case analyses showed smaller E-O associations, no evidence of M-O associations for attendance of antenatal classes and centre-based childcare on 5y and 17y mental health, and in general, wider confidence intervals for other M-O associations (Table S8).

**Table 2.** Distribution of mediators by exposure status (Analytic sample)

| Characteristic | High, N = 2,668<sup>1</sup> | Middle, N = 10,037<sup>1</sup> | Low, N = 2,808<sup>1</sup> | p-value<sup>2</sup> |
| --- | --- | --- | --- | --- |
| **Neighbourhood pathway** |  |  |  |  |
| Neighbourhood class |  |  |  | <0.001 |
| Good conditions | 2,276 (85.3%) | 7,496 (74.7%) | 1,688 (60.1%) |  |
| Poor conditions | 392 (14.7%) | 2,541 (25.3%) | 1,120 (39.9%) |  |
| **Services and social support pathway** |  |  |  |  |
| Attended antenatal classes |  |  |  | <0.001 |
| No | 1,194 (46.5%) | 5,995 (63.3%) | 2,126 (82.8%) |  |
| Yes | 1,373 (53.5%) | 3,474 (36.7%) | 443 (17.2%) |  |
| Attended 6 months centre-based childcare by 32 months |  |  |  | <0.001 |
| No | 1,624 (65.5%) | 7,623 (84.7%) | 2,184 (95.1%) |  |
| Yes | 856 (34.5%) | 1,379 (15.3%) | 113 (4.9%) |  |
| Low social support |  |  |  | <0.001 |
| No | 2,395 (93.4%) | 9,464 (96.8%) | 2,406 (93.4%) |  |
| Yes | 169 (6.6%) | 314 (3.2%) | 169 (6.6%) |  |
| **Material pathway** |  |  |  |  |
| Material class |  |  |  | <0.001 |
| Good physical living conditions | 2,516 (94.3%) | 8,424 (83.9%) | 1,638 (58.3%) |  |
| Overcrowding with lower risk of poor housing conditions | 50 (1.9%) | 615 (6.1%) | 588 (20.9%) |  |
| Poor housing conditions with lower risk of overcrowding | 102 (3.8%) | 998 (9.9%) | 582 (20.7%) |  |
| **Household adversity pathway** |  |  |  |  |
| Domestic violence |  |  |  | <0.001 |
| No | 2,167 (83.9%) | 6,980 (80.8%) | 1,556 (79.5%) |  |
| Yes | 417 (16.1%) | 1,662 (19.2%) | 402 (20.5%) |  |
| Substance misuse |  |  |  | <0.001 |
| No | 2,279 (93.1%) | 7,916 (91.0%) | 1,648 (89.7%) |  |
| Yes | 168 (6.9%) | 781 (9.0%) | 189 (10.3%) |  |
| Maternal mental distress | 2.7 (2.7) | 3.2 (3.7) | 4.4 (4.7) | <0.001 |
| **Parent-child relations pathway** |  |  |  |  |
| Disciplinary practices | 13.2 (4.5) | 13.1 (5.0) | 12.2 (5.5) | <0.001 |
| Parent-child relationship | 50.3 (6.2) | 49.3 (7.0) | 46.7 (7.9) | <0.001 |
| Parenting activities | 33.3 (7.1) | 31.3 (7.5) | 28.0 (8.5) | <0.001 |
| <sup>1</sup> n (%); Mean (SD) |  |  |  |  |
| <sup>2</sup> Pearson's Chi-squared test; One-way ANOVA |  |  |  |  |

**Table 2.** Associations between Exposure-Outcome and Mediator-Outcome, separately adjusted for each exposure and mediator variable. Multiply imputed data (m=10) with pooled estimates.

| Predictor | Parent-reported mental health 5y (n=15513) | Parent-reported mental health 17y (n=15513) |
| --- | --- | --- |
| **Childhood socioeconomic circumstances [Ref: High]** |  |  |
| Middle | 1.25 [1.04;1.46] *** | 1.44 [1.17;1.71] *** |
| Low | 3.33 [3.06;3.61] *** | 3.13 [2.78;3.49] *** |
| **Neighbourhood pathway** |  |  |
| Neighbourhood class [Ref: Good conditions] |  |  |
| Poor conditions | 1.28 [1.1;1.46] *** | 1.33 [1.06;1.6] *** |
| **Early support pathway** |  |  |
| Attended antenatal classes [Ref: No] | -0.24 [-0.43;-0.05] * | -0.35 [-0.63;-0.08] * |
| Attended centre-based childcare [Ref: No] | -0.23 [-0.46;0.01] | -0.31 [-0.63;0.02] |
| Low social support [Ref: No] | 0.47 [0.04;0.89] * | 0.71 [0.08;1.33] * |
| **Home environment pathway** |  |  |
| Material class [Ref: Good physical conditions] |  |  |
| Overcrowding with lower risk of poor housing conditions | 0.88 [0.56;1.21] *** | 1.2 [0.73;1.66] *** |
| Poor housing conditions with lower risk of overcrowding | 1.07 [0.79;1.35] *** | 1.2 [0.8;1.6] *** |
| **Household adversity pathway** |  |  |
| Domestic violence [Ref: No] | 0.64 [0.41;0.86] *** | 0.77 [0.41;1.13] *** |
| Substance misuse [Ref: No] | 0.22 [-0.06;0.51] | 0.17 [-0.26;0.6] |
| Maternal mental distress | 0.30 [0.27;0.32] *** | 0.23 [0.19;0.27] *** |
| **Parent-child relational pathway** |  |  |
| Disciplinary practices | 0.16 [0.14;0.18] *** | 0.09 [0.05;0.12] *** |
| Parent-child relationship | -0.26 [-0.27;-0.25] *** | -0.17 [-0.19;-0.14] *** |
| Parenting activities | -0.07 [-0.09;-0.06] *** | -0.04 [-0.06;-0.02] *** |

Note: Each exposure and mediating variable was estimated separately with the following adjustment sets: *Exposure: sex, ethnicity, maternal age, maternal mental health history, maternal childhood adversity (care experience)* *Mediators: additionally socioeconomic circumstances, all mediators in an earlier pathway, children in household, language spoken at home, gestational age, early developmental delays, long-term-limiting illness, parental high-frequency alcohol use* *Mediator-Outcome associations are not adjusted for mediators in the same pathway.*

### Interventional Disparity Measures

The total association between Low versus High SEC and 5y and 17y parent-reported mental health obtained in Monte Carlo simulations was around 3 points. Intervention on all five pathways *en bloc* gave indirect effects of −1.02[−1.32; −0.73] and −1.11[−1.54; −0.88] points, and residual associations of 2.22[1.93; 2.61] and 2.04[1.59; 2.54] points. The relative reduction in the total association was around one-third, at both ages. (See Table 3 and Figure S4). Point estimates from preliminary complete case analyses under-estimated the absolute size of the AdjTA, but showed the same relative reduction in the AdjTA of around a third (Table S2).

**Table 3.** Interventional Disparity Measures, Monte Carlo simulations with multiply imputed data (n=3,102,600 Monte Carlo observations)

|  | Parent-reported mental health 5y |  | Parent-reported mental health 17y |  |
| --- | --- | --- | --- | --- |
|  | Score difference (Low vs High SEC) | % Total association | Score difference (Low vs High SEC) | % Total association |
| **IDM** |  |  |  |  |
| Total association (AdjTA) | 3.24 [2.99;3.61] | 100% | 3.15 [2.87;3.68] | 100% |
| Indirect effect (IE) | -1.02 [-1.32;-0.73] | -32% [-40;-23] | -1.11 [-1.54;-0.88] | -35% [-48;-26] |
| Residual association (RA) | 2.22 [1.93;2.61] | 68% [60;77] | 2.04 [1.59;2.54] | 65% [52;74] |
| **Indirect effect via five early childhood pathways** |  |  |  |  |
| Neighbourhood | -0.14 [-0.22;-0.06] | -4% [-7;-2] | -0.07 [-0.13;0.06] | -2% [-4;2] |
| Social support | -0.02 [-0.24;0.17] | -1% [-8;5] | -0.33 [-0.67;-0.2] | -11% [-21;-6] |
| Material home environment | -0.09 [-0.18;0.02] | -3% [-6;1] | -0.16 [-0.27;-0.02] | -5% [-9;0] |
| Household adversity | -0.06 [-0.1;0.01] | -2% [-3;0] | -0.06 [-0.13;0] | -2% [-4;0] |
| Parent-child relational | -0.72 [-0.9;-0.54] | -22% [-27;-16] | -0.47 [-0.71;-0.31] | -15% [-22;-9] |
| Mediated dependence | 0.01 [-0.05;0.04] | 0% [-2;1] | 0 [-0.06;0.02] | 0% [-2;1] |

For 5y parent-reported total difficulties, the greatest reduction in the total association by intervention on early childhood mediators was predominantly attributable to the parent-child relational pathway (−22%[−27;−16]), with smaller contributions of the neighbourhood pathway (−4%[−7;−2]). There was weak evidence for small contributions of the material home environment (−3%[−6;1]), and household adversity pathways (−2%[−3;0]) and no evidence for contributions of the social support pathway (−1%[−8;5]), or mediated dependence (0%[−2;1]). For 17y parent-reported difficulties, the parent-child relational pathway (−15%[−22;−9]), and Social Support pathway (−11%[−21;−6]) were most important, with smaller contributions of the home environment (−5%[−9;0]) and household adversity (−2%[−4;0]) pathway. There was no evidence for a contribution of the neighbourhood pathway (−2%[−4;2]) or mediated dependence (0% [−2;1]). (See Table 3 and Figure 3). All point-estimates in 5y and 17y complete case analyses were within the 95% confidence intervals of the estimates in the main analysis (Table S2).

**Figure 3.**
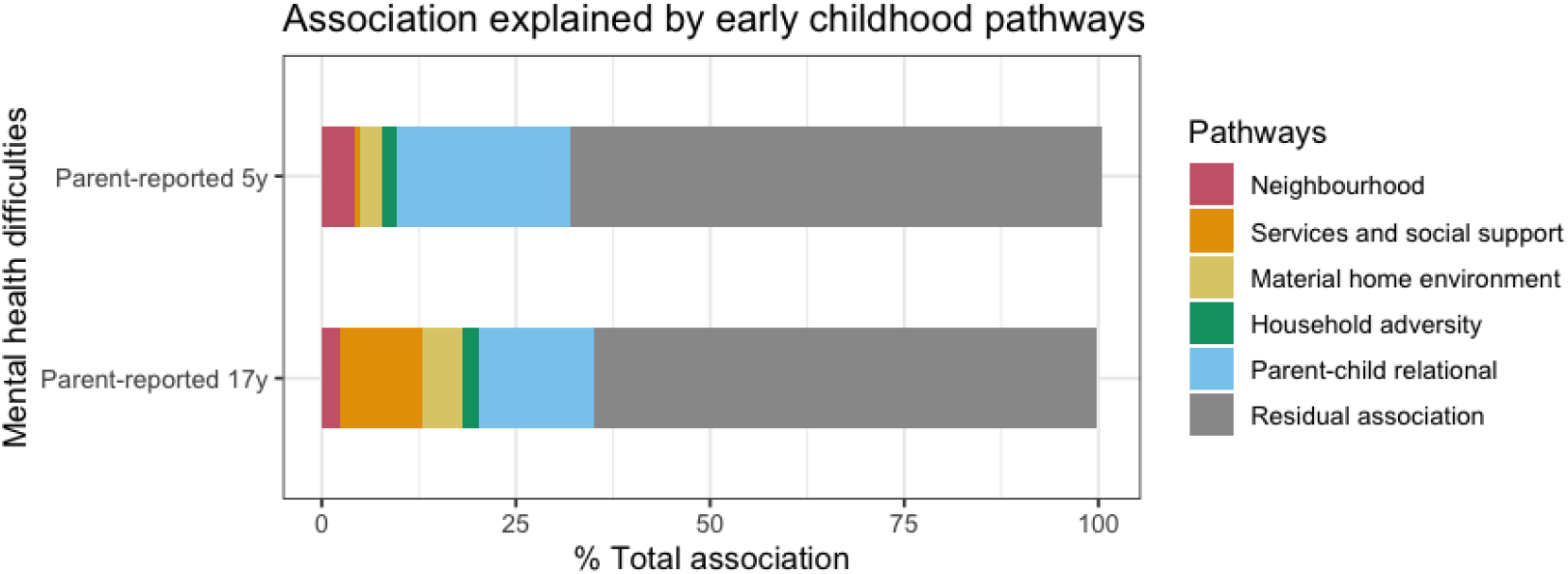
Decomposition of the percentage of the total association explained by mediators in five early childhood pathways. Monte Carlo simulations with multiply imputed data. Mediated dependence was 0%, so not depicted.

### Sensitivity analyses

For 17y self-reported total difficulties, we estimated a confounder-adjusted difference of 1.39[1.03;1.75] points, indicating greater self-reported difficulties in the Low vs High SEC group (Table S9). In Monte Carlo simulations, we estimated a similar adjusted total association of 1.49[1.27;2.03] points, with an indirect effect of −0.63[−1.07;−0.56] points, and a residual association of 0.86[0.40;1.37] points. The relative reduction in the total association due to intervention on early childhood mediators was −42%[−73;−20]. The social support pathway was most important (−21.6%[−46.7;−7.4]), followed by weak evidence of smaller contributions of the neighbourhood (−8%[−15;1]) and parent-child relational pathways (−8%[−26;3]), and no evidence for contributions of the home environment (−4%[−12;8]), household adversity (−1%[−5;5]) or mediated dependence (0%[−2;3]) (Table S10 and Figure S5)

At 17y, we estimated an odds ratio of 2.21[1.65;2.97], indicating around 2 times greater odds of meeting SDQ screening criteria for a probable mental disorder in the Low compared to High SEC group (Table S9). The total association, i.e., adjusted-percentage-point difference in meeting SDQ screening criteria was 9.1[7.2;11.1] percentage-points (Table S10). Intervening on the early childhood environment could reduce the difference in the absolute prevalence of a psychiatric disorder in the Low relative to High SEC group by 4.2[−6.2; −3.5] percentage-points, which is a −47%[−69;−35] relative change. The parent-child relational pathway (−19%[−28;−8]), social support (−17%[−39;−12]) were most important, with smaller contributions of the home environment (−9%[−17;−1]), no evidence for contribution of the neighbourhood (−1%[−5;7]) adversity pathway (−1%[−4;3]), or mediated dependence (0%[−3;3]) (Table S10 and Figure S6)

## Discussion

Socioeconomic inequalities in mental health difficulties at 5 and 17 years in the UK could be reduced by around one-third through hypothetical interventions in all major aspects of the early childhood environment. The parent-child relations pathway was the most influential, explaining over 20% of inequalities at 5 years. At 17 years, both early parent-child relations and social support networks play major roles, with social support especially important when considering adolescents’ own reports of their mental health. Intervening on the neighbourhood, home environment or household adversity may further reduce mental health inequalities, but to a lesser extent unless they also improve social support and parent-child relations. To reduce inequalities in child and adolescent mental health, neighbourhood-based approaches, improvements to the home environment and interventions addressing household adversity should be underpinned by strengthened community social support networks for parents, carers and children, alongside the promotion of positive parent-child relationships.

The estimated reduction of mental health inequality – between 25% to 50% as indicated by the 95% confidence intervals – is consistent with previous research despite methodological differences in the definitions of pathways and analytical approaches (16,17). Previous studies similarly emphasise the importance of parenting for shaping mental health inequalities, although they have often grouped parenting behaviours together with parental mental health (16,17). The importance of positive parent–child interactions and a stimulating home environment for later mental health is well established. Parents’ play a role in creating a stimulating home learning environment and in co-regulation of emotional stress. This supports the development early socioemotional and cognitive abilities (32–34), abilities needed to adapt and adjust well to new challenges later in adolescence. Especially in adolescence, negative dynamics in parent-child communication can influence an individual’s mental health by triggering conflict, impacting developing self-esteem and abilities to express emotions adaptively (35). The quality of parent-child relationships continue to have an impact on adulthood mental health in the UK, even when most adults live independently (36), supporting a lifelong role of positive early relationships in enabling emotional and social support when individuals navigate challenges in life.

While some earlier studies suggest that parental mental health and exposure to other adverse childhood experiences are central drivers of inequalities (8,37), we find that the adversity pathway accounts for little independent associations once parent–child relations are taken into account. This difference could be because our parenting pathway includes some variables capturing disciplinary practices - previously used to define child maltreatment as an adverse childhood experience (8). Our findings could also indicate the importance of focusing on whether children have safe and supporting relationships at home even in the presence of adversity (38), and supporting positive parenting, which has protective effects independent of adversity (39). In line with this, family emotional support (40) and parent-child communication (41) has been shown to mitigate detrimental effects of adversity on child and adolescent mental health.

Contrary to some prior findings (17), we do not identify an independent contribution of neighbourhood factors to adolescent mental health inequalities once other pathways are adjusted for. Earlier evidence also suggest that universal centre-based childcare, in this study considered under the social support pathway, can reduce inequalities, but this may reflect benefits that depend on universal uptake (42) or benefits specific to externalising difficulties (9). Furthermore, in this study, neighbourhood factors and childcare access are considered simultaneously alongside several other mediator pathways, which may attenuate the independent effect of neighbourhood and social support.

However, the role of disciplinary practices requires further clarification to differentiate harmful from supportive discipline. Our measure combines hostile disciplinary practices (such as shouting or smacking) with boundary-setting strategies (such as taking away treats) (8). Hostile practices are known to undermine children’s mental health (43,44), while consistent boundaries can support positive development (45). Individuals in the high-SEC group tended to use more disciplinary strategies overall – therefore, the simulated hypothetical intervention shifted the distribution of disciplinary strategies in the low-SEC group towards greater use of such practices. As a result, we may have underestimated the contribution of the parenting pathway to mental health inequalities due to inequalities in harmful disciplinary practices.

Our findings carry implications for policy and practice. Mental health services in the UK are under considerable strain, with resources concentrated heavily on crisis management or targeted school-based interventions often for older children. Yet, we show here that tackling mental health inequalities needs to start before school age. Potential interventions could include increasing access to evidence-based parenting programmes of which several are already available in the UK (46,47), policies that expand access to high-quality childcare (48), and increase efforts to mitigate material and environmental stressors on parent-child relationships. Parenting interventions also need to address how confident the parent feels in establishing a relationship with their child – influenced by parental self-esteem and mental health, alongside their own experiences as a child. Services will also need to address barriers to uptake and engagement with interventions especially in disadvantaged groups, to maximise the benefits on inequality reduction (49). Evaluations of early childhood interventions could measure benefits on access to social support and parent-child relations, as they are key factors for addressing mental health inequalities.

This study also highlights the need for systems-level reforms, workforce development, and better integration of services supporting families. The early years offer universal touchpoints, such as early education settings and mandated health visitor appointments in the UK (50), which could be leveraged more effectively to support children and parents. However, very often these sectors operate with differing priorities driving silo working. Health visitors face competing demands to monitor child development, safeguarding and as well as increasingly complex caseloads that can affect their capacity to support families effectively, whilst early years settings focus primarily on academic preparation rather than family support. While families interact with different services for a specific need (e.g., speech and language, physical therapy), these services are often disconnected. Systems which enable professionals working with young children to collaborate, to improve early identification of needs and provide support for challenges faced at home will enable children to thrive (51). In England the new integrated family hubs model (52), jointly overseen by the Department for Health and Social Care and the Department for Education is a promising approach to foster cross-sector working, reduce fragmentation and improve equity of access for early support. Family Hubs are also expected to provide a vital link to improve connectedness to the local community and support networks, improve access to health knowledge and therefore, empower families facing adversity and families with children facing developmental needs.

### Strengths, limitations and future directions

A key strength of this study is the use of interventional disparity measures (IDMs), which rely on fewer causal assumptions, i.e., the absence of mediator-outcome confounding, alongside consistency and no interference (further discussed in Appendix S4). Since we considered a large set of environmental mediators alongside early life confounders, there are few unmeasured factors that can still influence the mediator-outcome relationships. However, any unmeasured confounders and mediator pathways that have not been considered may still bias our results. IDMs are also robust to different assumptions about the ordering of mediators – i.e., even if the true ordering of mediators were different to our proposed distal to proximal order, the estimated effects via each mediator pathway remains the same (53). We considered a wide range of environmental factors organised into five pathways, developed through collaboration with public child health practitioners. This ensures that the findings are relevant to real-world applications and provide rapid insight into which broad areas may be most promising for reducing inequalities.

However, the broad scope of mediators also presents limitations. Grouping many variables into pathways makes it difficult to disentangle the specific mechanisms operating within each pathway or to fully characterise the complexity within each domain. For example, data reduction may have limited the variability captured within the material and neighbourhood pathways. The theoretical approach also limited a detailed characterisation of the role of early childhood education, which falls under the social support pathway in our conceptualisation but is itself, presently, a key focus of early years policy worldwide. Healthcare access was measured only through antenatal care attendance due to limited data. Considering a large set of mediators also results in high computational time and greater missing data in complete case analyses. We addressed missing data using imputation which produced similar findings to complete-case analyses.

We estimated the reduction in inequality between groups differing in maternal education, because it is relatively fixed from birth. However, alternative measures of socioeconomic disadvantage – such as household income, family structure, or neighbourhood deprivation – could emphasise different pathways or mechanisms (54).

Additionally, because the study uses observational data, our findings reflect the potential effects of hypothetical interventions rather than the proven effectiveness of specific programmes.

Future research should focus on clarifying the relationships between pathways and identifying modifying factors. Priorities include understanding how adversity influences parenting in disadvantaged families, how neighbourhood conditions affect access to social support, and how child characteristics—such as sex—shape the influence of parenting styles (55). Unpacking the importance of variables within pathways (for example, distinguishing the access to early childhood education from other forms of social support) will help refine policy recommendations. Comparing the role of early childhood and adolescent pathways could also contribute valuable insight. This is because adolescence provides another important window for intervening on emerging mental health problems. An individual’s socioeconomic circumstances expand to include peers and schools, and this could require different interventions such as through schools, youth workers and access to mental health interventions, which are different to the intervention types implicated in this analysis.

## Conclusions

In conclusion, hypothetical interventions on the early childhood environment have the potential to reduce socioeconomic inequalities in child and adolescent mental health by a third. Interventions on parent–child relationships and social support can lead to the largest inequality reduction. However, interventions must address the broader socioeconomic and contextual factors that shape individual behaviours related to social support and parent-child relationships.

## Acknowledgements

We thank the families taking part in the Millennium Cohort Study for their time and commitment to the study, and research team carrying out the Millennium Cohort Study at the UCL Centre for Longitudinal Studies.

## Funding

YWC is part-funded by an NIHR Research Professorship (NIHR302438) awarded to DTR, and by NIHR Oxford Health Biomedical Research Centre. DTR is funded on an NIHR Research Professorship (NIHR302438) and the NIHR School for Public Health Research (grant reference number NIHR204000). VSS is supported by the Swedish Research Council for Health, Working Life and Welfare (FORTE 2024-00264). ETCL is supported by the CUHK Vice-Chancellor’s One-off Discretionary Fund (grant number: 136604080) and the Hong Kong Jockey Club Charity Trusts (grant number 2024-0058-003). The views expressed are those of the authors and not necessarily those of the NIHR or the Department of Health and Social Care. For the purpose of open access, the author has applied a Creative Commons Attribution (CC BY) licence to any Author Accepted Manuscript version arising from this submission.

## Declarations

### Author contributions

Conceptualisation: Yu Wei Chua, David Taylor-Robinson; Methodology: Yu Wei Chua, Philip McHale, Daniela Schlüter, Viviane S. Straatmann, Eric T.C. Lai, Melisa Campbell, Bethan Plant, Michelle Black, David Taylor-Robinson; Data curation: Yu Wei Chua; Formal analysis: Yu Wei Chua; Writing – original draft: Yu Wei Chua;

Writing – review and editing: Yu Wei Chua, Philip McHale, Viviane S. Straatmann, Eric T.C. Lai, Melisa Campbell, Bethan Plant, Michelle Black, David Taylor-Robinson

### Competing interests

The authors have no competing interests to declare.

### Ethics approval

Ethical approval for the Millennium Cohort Study (MCS) was provided by an NHS Research Ethics Committee, in accordance with the 1964 Declaration of Helsinki and its later amendments or comparable ethical standards. No further ethical approval was required for this secondary analysis of MCS data.

## Data and code availability

Data from the UK Millennium Cohort Study is openly available on the UK Data Service (DOI: https://doi.org/10.5255/UKDA-Series-2000031). R code for processing MCS data, descriptive statistics, and simulating IDMs will be made openly available via the Open Science Framework (OSF) upon publication.

## Notes

### Competing Interest Statement

The authors have declared no competing interest.

### Author Declarations

https://doi.org/10.5255/UKDA-Series-2000031

## References

1. Reiss F. Socioeconomic inequalities and mental health problems in children and adolescents: a systematic review. Soc Sci Med. 2013 Aug;90:24–31.

2. Cadman T, Avraam D, Carson J, Elhakeem A, Grote V, Guerlich K, et al. Social inequalities in child mental health trajectories: a longitudinal study using birth cohort data 12 countries. BMC Public Health. 2024 Oct 22;24(1):2930.

3. Chua YW, Schlüter D, Pearce A, Sharp H, Taylor-Robinson D. Socioeconomic inequalities in mental health difficulties over childhood: a longitudinal sex-stratified analysis using the UK Millennium Cohort Study. Soc Sci Med. 2025 May 4;378:118159.

4. Putra IGNE, McInerney AM, Robinson E, Deschênes SS. Neighbourhood characteristics and socioeconomic inequalities in child mental health: Cross-sectional and longitudinal findings from the Growing Up in Ireland study. Health & Place. 2024 Mar 1;86:103180.

5. Rutherford C, Sharp H, Hill J, Pickles A, Taylor-Robinson D. How does perinatal maternal mental health explain early social inequalities in child behavioural and emotional problems? Findings from the Wirral Child Health and Development Study. PLOS ONE. 2019 May 24;14(5):e0217342.

6. Verhulst F, Tiemeier H. Socioeconomic Inequalities and Mental Health Problems in Children and Adolescents. In: Taylor E, Verhulst F, Wong JCM, Yoshida K, editors. Mental Health and Illness of Children and Adolescents [Internet]. Singapore: Springer; 2020 [cited 2025 Jan 16]. p. 257–74. Available from: 10.1007/978-981-10-2348-4_57

7. McLaughlin KA, Breslau J, Green JG, Lakoma MD, Sampson NA, Zaslavsky AM, et al. Childhood socio-economic status and the onset, persistence, and severity of DSM-IV mental disorders in a US national sample. Social Science & Medicine. 2011 Oct 1;73(7):1088–96.

8. Straatmann VS, Lai E, Law C, Whitehead M, Strandberg-Larsen K, Taylor-Robinson D. How do early-life adverse childhood experiences mediate the relationship between childhood socioeconomic conditions and adolescent health outcomes in the UK? J Epidemiol Community Health. 2020 Nov 1;74(11):969–75.

9. Green MJ, Pearce A, Parkes A, Robertson E, Katikireddi SV. Pre-school childcare and inequalities in child development. SSM - Population Health. 2021 June 1;14:100776.

10. Robertson E, Leyland A, Pearce A. The potential of early years’ childcare to reduce mental health inequalities of school age children in Scotland. SSM - Population Health. 2024 June 1;26:101682.

11. Kelly Y, Sacker A, Bono ED, Francesconi M, Marmot M. What role for the home learning environment and parenting in reducing the socioeconomic gradient in child development? Findings from the Millennium Cohort Study. Archives of Disease in Childhood. 2011 Sept 1;96(9):832–7.

12. Tamura K, Morrison J, Pikhart H. Children’s behavioural problems and its associations with socioeconomic position and early parenting environment: findings from the UK Millennium Cohort Study. Epidemiology and Psychiatric Sciences. 2020 Jan;29:e155.

13. Pearce A, Dundas R, Whitehead M, Taylor-Robinson D. Pathways to inequalities in child health. Arch Dis Child. 2019 Feb 23;

14. Link BG, Phelan J. Social conditions as fundamental causes of disease. J Health Soc Behav. 1995;Spec No:80–94.

15. Pearce A, Hope S, Green MJ, Lynch JW, Oude Groeniger J, De Stavola B, et al. Causal mediation approaches for understanding pathways to inequalities and policy entry points: examples from early years health and development. J Epidemiol Community Health. 2025 Nov 3;jech-2025-224260.

16. Cattan S, Fitzsimons E, Goodman A, Ploubidis G, Phimister A, Wertz J. Early childhood inequalities [Internet]. The IFS; 2022 June [cited 2023 July 24]. Available from: http://default/publications/early-childhood-inequalities-0

17. Straatmann VS, Lai E, Lange T, Campbell MC, Wickham S, Andersen AMN, et al. How do early-life factors explain social inequalities in adolescent mental health? Findings from the UK Millennium Cohort Study. J Epidemiol Community Health. 2019 Nov 1;73(11):1049–60.

18. Geelhoed EA, Bloom DE, Bock C, Flatau P, Mandzufas J, Li I, et al. Informing Resource Allocation for Investment in Early Childhood: A Review of the International Peer-Reviewed Evidence. Australian Economic Review. 2022;55(2):215–31.

19. Tanner J, Candland T, Odden WS. Later Impacts of Early Childhood Interventions: A Systematic Review | Independent Evaluation Group [Internet]. Independent Evaluation Group, The World Bank Group; 2015 [cited 2024 July 25]. Available from: https://ieg.worldbankgroup.org/evaluations/later-impacts-early-childhood-interventions

20. Jeong J, Franchett EE, Oliveira CVR de, Rehmani K, Yousafzai AK. Parenting interventions to promote early child development in the first three years of life: A global systematic review and meta-analysis. PLOS Medicine. 2021 May 10;18(5):e1003602.

21. Shaundreal J. Educational and Health Benefits of Early Childhood Education Interventions for Low-Income Children: A Systematic Review. 2020;

22. Dahlberg M, Nordmyr J, Gunnarsdottir H, Forsman AK. The Evidenced Effects of Early Childhood Interventions to Promote Mental Health and Parenting in the Nordic Countries: A Systematic Review. IJMHP. 2023;25(4):505–37.

23. Morrison J, Pikhart H, Ruiz M, Goldblatt P. Systematic review of parenting interventions in European countries aiming to reduce social inequalities in children’s health and development. BMC Public Health. 2014 Oct 6;14(1):1040.

24. Fernainy P, Cohen AA, Murray E, Losina E, Lamontagne F, Sourial N. Rethinking the pros and cons of randomized controlled trials and observational studies in the era of big data and advanced methods: a panel discussion. BMC Proceedings. 2024 Jan 18;18(2):1.

25. Connelly R, Platt L. Cohort Profile: UK Millennium Cohort Study (MCS). International Journal of Epidemiology. 2014 Dec 1;43(6):1719–25.

26. Goodman R. Psychometric Properties of the Strengths and Difficulties Questionnaire. Journal of the American Academy of Child & Adolescent Psychiatry. 2001 Nov 1;40(11):1337–45.

27. OECD. What are the benefits of ISCED 2011 classification for indicators on education? [Internet]. 2015 Nov [cited 2024 July 24]. Available from: https://www.oecd-ilibrary.org/education/what-are-the-benefits-of-isced-2011-classification-for-indicators-on-education_5jrqgdw9k1lr-en

28. United Nations Educational, Scientific and Cultural Organisation (UNESCO). International Standard Classification of education: ISCED 2011 [Internet]. UNESCO Institute for Statistics; 2012 [cited 2024 July 24]. Available from: https://uis.unesco.org/sites/default/files/documents/international-standard-classification-of-education-isced-2011-en.pdf

29. Centre for Longtudinal Studies. Millennium Cohort Study: User Guide (Surveys 1 – 5) 9th Edition. London: UCL Institute of Education; 2020 Aug.

30. Micali N, Daniel RM, Ploubidis GB, De Stavola BL. Maternal Prepregnancy Weight Status and Adolescent Eating Disorder Behaviors: A Longitudinal Study of Risk Pathways. Epidemiology. 2018 July;29(4):579– 89.

31. McHale P, Schlüter DK, Abbasizanjani H, Akbari A, Barr B, Taylor-Robinson D. Mediation of socioeconomic inequalities in preterm birth. A cohort analysis of Welsh linked data. Acta Obstetricia et Gynecologica Scandinavica. 2025;104(6):1081–91.

32. Kelly Y, Sacker A, Bono ED, Francesconi M, Marmot M. What role for the home learning environment and parenting in reducing the socioeconomic gradient in child development? Findings from the Millennium Cohort Study. 2011 Sept 1 [cited 2026 Jan 2]; Available from: https://adc.bmj.com/content/96/9/832.short

33. Liang Y, Cao H, Zhou N, Li J, Zhang L. Early home learning environment predicts early adolescents’ adjustment through cognitive abilities in middle childhood. Journal of Family Psychology. 2020;34(8):905– 17.

34. Paley B, Hajal NJ. Conceptualizing Emotion Regulation and Coregulation as Family-Level Phenomena. Clin Child Fam Psychol Rev. 2022 Mar 1;25(1):19–43.

35. Zapf H, Boettcher J, Haukeland Y, Orm S, Coslar S, Fjermestad K. A systematic review of the association between parent-child communication and adolescent mental health. JCPP Advances. 2024;4(2):e12205.

36. Morgan Z, Brugha T, Fryers T, Stewart-Brown S. The effects of parent–child relationships on later life mental health status in two national birth cohorts. Soc Psychiatry Psychiatr Epidemiol. 2012 Nov 1;47(11):1707–15.

37. Lai ET, Schlüter DK, Lange T, Straatmann V, Andersen AMN, Strandberg-Larsen K, et al. Understanding pathways to inequalities in child mental health: a counterfactual mediation analysis in two national birth cohorts in the UK and Denmark. BMJ Open. 2020 Oct 12;10(10):e040056.

38. Dooley B, Fitzgerald A, Giollabhui NM. The risk and protective factors associated with depression and anxiety in a national sample of Irish adolescents. Ir j psychol Med. 2015 Mar;32(1):93–105.

39. Yamaoka Y, Bard DE. Positive Parenting Matters in the Face of Early Adversity. American Journal of Preventive Medicine. 2019 Apr;56(4):530–9.

40. Adjei NK, Jonsson KR, Straatmann VS, Bellavia A, Yaya S, McGovern R, et al. Perceived emotional support mediates the association between childhood family adversity and adolescent mental health in the UK millennium cohort. Sci Rep. 2025 Nov 28;15(1):42730.

41. Ying L, Zhou H, Yu S, Chen C, Jia X, Wang Y, et al. Parent–child communication and self?esteem mediate the relationship between interparental conflict and children’s depressive symptoms. Child. 2018 Nov;44(6):908–15.

42. Barry KM, Avraam D, Cadman T, Elhakeem A, Marroun HE, Jansen PW, et al. Early childcare arrangements and children’s internalizing and externalizing symptoms: an individual participant data meta-analysis of six prospective birth cohorts in Europe. The Lancet Regional Health – Europe [Internet]. 2024 Oct 1 [cited 2025 Oct 28];45. Available from: https://www.thelancet.com/journals/lanepe/article/PIIS2666-7762(24)00203-5/fulltext

43. Kingsbury M, Sucha E, Manion I, Gilman SE, Colman I. Adolescent Mental Health Following Exposure to Positive and Harsh Parenting in Childhood. Can J Psychiatry. 2020 June;65(6):392–400.

44. Katsantonis I, Symonds JE. Population heterogeneity in developmental trajectories of internalising and externalising mental health symptoms in childhood: differential effects of parenting styles. Epidemiology and Psychiatric Sciences. 2023 Jan;32:e16.

45. Wong TKY, Konishi C, Kong X. Parenting and prosocial behaviors: A meta-analysis. Social Development. 2021;30(2):343–73.

46. Han MX, Chesney E, Ng V, Bright J, Sagar YK, Baker E, et al. Universal, selective and indicated parenting interventions to prevent the development of adverse mental health outcomes in youth: a meta-review of systematic reviews. BMJ Ment Health [Internet]. 2025 July 3 [cited 2025 Oct 28];28(1). Available from: https://mentalhealth.bmj.com/content/28/1/e301613

47. Foundations. Effective interventions and practices for parents experiencing complex and multiple needs - Foundations [Internet]. 2025 [cited 2025 Oct 28]. Available from: https://foundations.org.uk/our-work/current-projects/effective-interventions-and-practices-for-parents-experiencing-complex-and-multiple-needs/

48. Hobbs A, Bernard R. Impact of COVID-19 on Early Childhood Education & Care. 2021 Oct 27 [cited 2025 Jan 22]; Available from: https://post.parliament.uk/impact-of-covid-19-on-early-childhood-education-care/

49. Lowther-Payne HJ, Ushakova A, Beckwith A, Liberty C, Edge R, Lobban F. Understanding inequalities in access to adult mental health services in the UK: a systematic mapping review. BMC Health Serv Res. 2023 Sept 29;23(1):1042.

50. Office for Health Improvement and Disparities. Healthy child programme: health visitor and school nurse commissioning [Internet]. 2016 [cited 2024 Nov 14]. Available from: https://www.gov.uk/government/collections/healthy-child-programme

51. World Health Organisation, United Nations Children’s Fund, World Bank Group. Nurturing care for early childhood development: a framework for helping children survive and thrive to transform health and human potential [Internet]. Geneva; 2018 [cited 2025 Dec 30]. Available from: https://www.who.int/publications/i/item/9789241514064

52. HM Government. Family Hubs and Start for Life programme guide 2025–26 [Internet]. 2025. Available from: https://assets.publishing.service.gov.uk/media/67cacd6ba175f08d198d80c1/Family_Hubs_and_Start_for_Life_programme_guide_2025-26.pdf

53. Vansteelandt S, Daniel RM. Interventional Effects for Mediation Analysis with Multiple Mediators. Epidemiology. 2017 Mar;28(2):258.

54. Galobardes B, Shaw M, Lawlor DA, Lynch JW, Smith GD. Indicators of socioeconomic position (part 1). Journal of Epidemiology & Community Health. 2006 Jan 1;60(1):7–12.

55. Kingsbury M, Sucha E, Manion I, Gilman SE, Colman I. Adolescent Mental Health Following Exposure to Positive and Harsh Parenting in Childhood. Can J Psychiatry. 2020 June;65(6):392–400.

