## Supplementary Information for "Reducing mental health inequalities through hypothetical interventions in early childhood: Evidence from mediation analysis using the UK Millennium Cohort Study"

### Contents

S1. Additional methodological notes
S2. Operationalisation of mediating variables in the Millennium Cohort Study (MCS)
S3. Operationalisation of covariates in the Millennium Cohort Study (MCS)
S4. Interventional Disparity Measures
S5. Interventional Disparity Measures Definitions.
S6. Estimation by Monte Carlo simulation with imputation of missing data
S7. References
S8. Supplemental Figures and Tables

### S1. Additional methodological notes

#### *Selection of outcome*

Parent-report of the SDQ was used to assess meeting screening criteria for a psychiatric disorder as it has been shown to have a greater predictive value than self-report for both emotional and behavioural disorders (1). We only assessed presence of psychiatric disorder at 17 years (and not at 5 years) in sensitivity analyses due to the computational demand of Monte Carlo simulation, as continuous measures already capture full information in the SDQ measures and because the onset of most mental health disorders is in adolescence.

Adolescent self-report of mental ill health at 17 years on the Kessler-6 scale (2,3) is also available. This a measure of recent experience of general psychological distress, which is also predictive of severe mental illness. While it has good predictive value for mood and anxiety disorders, it has limited predictive value for behavioural disorders (4). Measurement non-invariance between males and females have also been reported, suggesting sex biases in self-report of psychological distress, and predictive utility for abnormal scores on the SDQ was low for males (5). As our focus was on any psychiatric disorder, and not to compare emotional and behavioural disorders, we did not consider this measure further given the computational demand of this simulation study.

#### *Categorisation of exposure*

We initially used all six maternal education categories (Degree plus, Diploma, A levels, GCSE A-C, GCSE D-G, None of these qualifications), but this affected model convergence due to the number of interactions between mediators and covariates considered, so we reduced it to a three-category variable.

#### *Overlap between definition of socioeconomic circumstances and early childhood environment mediators*

This approach explains mental health inequalities due to unequal childhood socioeconomic circumstances, by examining how risk and protective factors are unevenly distributed across socioeconomic groups. This study focuses on factors in the early childhood environment, defined as the early psychosocial and physical conditions children need for healthy development.

Socioeconomic circumstances are understood as multidimensional while the mediator pathways characterised by the early childhood environment are domain specific. For example, neighbourhood socioeconomic disadvantage (one measure of socioeconomic circumstances) is defined not only by residents' income but also by environmental conditions, service availability, housing quality, and social issues such as crime. While higher-income individuals may have more ability to move to better neighbourhoods, the neighbourhood pathway in this study focuses on neighbourhood conditions themselves rather than household wealth. Similarly, monetary or resource-based measures of socioeconomic circumstances, such as income and housing, reflect not just material resources but broader influences such as financial stress and neighbourhood quality, helping to characterise the material home environment more precisely. Therefore the material home environment pathway aims to capture more specifically, the physical characteristics of the home.

Different indicators of socioeconomic circumstances emphasise different dimensions of risk and protective factors. In other words, specific risk and protective pathways may be more important when considering groups that differ according to a socioeconomic indicator (e.g., education, income, occupation, housing tenure, family structure, or neighbourhood deprivation). However, similar patterns of socioeconomic characteristics often co-occur in the context of socioeconomic disadvantage (e.g., education influences income, education and family structure influences household income, greater income could lead to moving to a better-quality neighbourhood).

Rather than attempting to isolate the effect of a single socioeconomic factor, this study compares the most and least advantaged socioeconomic childhood socioeconomic circumstances, defined by maternal education level. As we do not adjust for other correlated socioeconomic dimensions, the study estimates the total inequality experienced in real-world conditions, where multiple disadvantages tend to co-occur.

#### *Participatory research consultation*

The initial groupings (neighbourhood, material, household adversity, parent-child relationship, educational environment) were discussed in a research consultation. The key takeaways were that the child's wider environment is predominantly in the home during the early years, but the neighbourhood may capture factors influencing parental health. Social support and engagement with early intervention services were missing from our initial framework, as the two are closely related; a lack of social support can mean that public services may be even more important. Although some parent-child activities can be considered educational, these are inextricable from formation of parent-child relationships, and access to childcare can also fall under a pathway to accessing early support. The consultation also stimulated later discussions within the research team about how the wider neighbourhood and social environment may be considered from a capabilities approach, a philosophy which focuses on ten central capabilities basic to human life which are needed to achieve wellbeing (6). Capabilities refer to the freedom of opportunity, or availability of means to carry out, choose and achieve value in domains of life, contrasting with a solely resource-based view. For example, the neighbourhood pathway maps onto the capability to have bodily integrity and engage in play, while the services and social support pathway maps in part, onto the capability to have affiliation with others.

***Complexity perspective***

Importantly, our approach aims to capture the difference in mental health difficulties experienced if we were able to intervene on the early childhood mediators, not to decompose the complex effects via early life, early childhood or downstream outcomes and later exposures. First, we hypothesise that within this period, more distal pathways can influence more proximal pathways, but our estimation method remains valid regardless of the underlying direction of the pathways. Although mediators are measured at both 9m and 3y, we do not aim to decompose the infancy and early childhood timing in which these variables act, but use these variables to capture the early childhood environment experienced around 9m and 3y.

Second, we take a complexity perspective on causal pathways (7). For example, these early childhood factors could influence subsequent child outcomes such as school readiness, that further influence later mental health outcomes. There can also be nonlinear effects where experience of a risk or protective factor leads to differential exposure and vulnerability to other risk and protective factors (e.g., the neighbourhood shaping the experience of later school factors such as bullying and experience of academic stress), as well as feedback mechanisms where downstream outcomes affect the levels of exposure to later risk or protective. Similarly, what we are interested in is the difference in mental health outcomes if we were able to intervene on these early life mediators, given the unknown nature of any complex downstream pathways.

***Data reduction approach***

We first carried out a data reduction approach to reduce the number of mediators in the neighbourhood and material pathway. We obtained mediator classes using Latent Class Analysis (*poLCA* in R) for binary Neighbourhood and Material indicators. We aimed to capture the most parsimonious and theoretically plausible groupings, and therefore only considered 1, 2, or 3 classes. Based on AIC, BIC, entropy and size of the smallest class (Table S1), we selected the 2-class model for the neighbourhood pathway, and 3-class model for the material pathway (Figure S1). We then re-ran the best-fitting model using Stata using the *svy: gsem* function which allowed for inclusion of survey weights to account for missing data bias from item non-response. We obtained the posterior probabilities for each identified class and each individual was assigned a class based on their most likely class.

***Model fitting***

Models of the outcome included all exposure, confounder and mediating variables shown in Figure 2. Marginal models of each mediator included exposure, baseline and intermediate confounders, and only the upstream mediators in the same pathway. Conditional models of each mediator included the exposure, baseline and intermediate confounders, all mediators in any preceding mediator pathway and upstream mediators in the same pathway. We estimated richly specified regression models, i.e., considering interactions between all variables included. Due to the large number of mediating variables, some models did not converge. As such, we proceeded by including interactions between exposure, mediators and covariates capturing the child's developmental characteristics (ie., sex, preterm birth, long-term limiting illness, early developmental delay) which are most likely to modify the effect child's environment on later mental health.

**S2. Definitions of mediating variables in the Millennium Cohort Study (MCS)**

| Variable | Age assessed | |
|---|---|---|
| ***Neighbourhood pathway*** | | |
| Environmental problems | 9 months | At 9 months, parents answered questions related to their neighbourhood living environment, access to services and crime.<br><br>Can you tell me how common each of these things is in your area? (1-Very common, Fairly common, not very common, 4-not at all common)<br><br>1. Rubbish or litter lying around<br>2. Vandalism and deliberate damage<br>3. Insults or attacks to do with someone's race or colour<br>4. Poor public transport<br>5. Food shops and supermarkets that are easy to get to?<br>6. Pollution, grime or other environmental problems<br><br>A question on religious intolerance was asked for MCS families in Northern Ireland (How common were insults or attacks to do with someone's religion). This item was not included.<br><br>Environmental problems (Yes, No) was defined as answering Very common or Fairly common to Item 1 and 6<br><br>Poor accessibility to services (Yes, No) was defined as answering Very common or Fairly common to Item 4 and Not very common or Not at all common to Item 5.<br><br>Problems with crime (Yes. No) was defined as answering Very common or Fairly common to Items 2 and 3. |
| Poor accessibility to services | 9 months | |
| Problems with crime | 9 months | |
| Places for children to play safely (Binary) | 9 months | At 9 months, parents were asked whether there were places in the neighbourhood for children to play safely *(Yes – safe places to play, No – no safe places to play).* |
| Living in a safe neighbourhood (Binary) | 3 years | At 3 years, parents were asked how safe they felt in the area they live in (Very safe, fairly safe, neither safe not unsafe, fairly unsafe, very unsafe). Living in a safe neighbourhood was defined as very or fairly safe. |
| ***Service engagement and social support pathway*** | | |
| Early engagement with healthcare services | 9 months | |
| Accessed 6 months centre-based childcare (binary) | 9 months<br>3 years | Main respondents were asked about main childcare arrangements at 9 months, and at 3 years, whether |

| | | |
|---|---|---|
| | | this had changed, and whether there were other childcare arrangements. For each childcare arrangement when it started, stopped or whether it is still going on.<br><br>Following previous work, we defined centre-based childcare as exposure to at least 6 months of centre-based childcare by 32 months which was the youngest age of follow up at the 3 years time point (8). |
| Low social support | 9 months | Main respondents were asked about contact with their parents, childcare arrangements and whether they have friends to offer support. Low social support was defined as having less than daily/weekly contact with mother and father, not having regular grandparental childcare, as well as not having any friends to offer support. |
| ***Material home environment*** | | |
| Can't afford essentials for child | 3 years | At 3 years, parents were asked about their standard of living, based on questions assessing material deprivation in the UK Family Resource Survey (9).<br><br>Do you have any of the following items (yes/no), and is this because you do not want this/these or cannot afford this/these (does not want/cannot afford)<br>1. A warm waterproof coat for child<br>2. New properly fitted shoes for child<br>3. Fresh fruit or vegetables at least once a day for child<br>4. Insurance for contents of your home<br>5. A small amount of money to spend on yourself weekly, not on the family<br>6. Holiday away from home once a year not staying with relatives<br>7. The ability to replace worn-out furniture<br><br>Material deprivation was defined as not being able to afford 3 or more of these items. Two other questions were also available at the 3 years follow up (Whether the parent has two pairs of weather-proof shoes for yourself, and whether parent has a hobby or leisure activity), but based on a review of the questions assessing material deprivation, these were not predominantly regarded as essential by UK families (10) |
| Problems with heating home | 9 months<br>3 years | At 9 months and 3 years, parents were asked about all the types of heating they used for their home (No heating (only one response allowed), central heating, coal fires, wood fires or stoves, gas fires, electric fires, paraffin heaters or other). Responses were converted into binary variables for each type of heating.<br><br>At 9 months, parents were asked about the temperature in the room that the cohort child sleeps in |

| | | |
|---|---|---|
| | | (very warm, warm, neither warm nor cold, cold, very cold)<br><br>Problems with heating the home was a binary variable defined as having no heating, no central heating at 9 months or 3 years, or if the baby's room was cold or very cold at 9 months. |
| Problems with damp | 3 years | At 9 months, parents were asked if there were ever any damp or condensation on the walls of their home (yes, no).<br><br>At 3 years, parents were asked if there were problems with damp condensation except in the kitchen or bathroom (no damp, not much of a problem, some problems or great problem)<br><br>Because the 9 month variable was not specific enough, we used the 3 year variable, where problems with damp was a binary variable defined as some problems or great problem with damp condensation apart from in the kitchen or bathroom. |
| Overcrowding | 9 months<br>3 years | At 9 months and 3 years, main respondent reported the number of rooms in the house and the total number of household members. We used the simplest measure of overcrowding, person-per-room, as most common overcrowding measures are valid in capturing material resources (Cable & Sacker, 2019).<br><br>Overcrowding was indicated if person-per-room>1 at either 9 months or 3 years, in other words number of household members exceed the number of rooms. We used both time points as overcrowding could occur later when younger siblings children are born. |
| Insecure housing | 9 months | At 9 months, parents were asked if, since cohort member was born whether there has ever been a time when they were homeless (had to move out of a place and had nowhere permanent to live) |
| Safe play environment at home | 3 years | At 3 years, interviewers visited participants' home to assess the child's home environment using selected items from the Home Observation and Measurement of the Environment-Short Form (HOME-SF) scale (Caldwell and Bradley, 1984)<br><br>An interviewer assessed if child's in-home environment was safe (Safe, Not safe, not observed), and whether parents provided toys during visit (Provided toys, Did not provide toys, not observed). If this was not observed, this variable was coded as missing. |
| Parent provided toys during visit | 3 years | |
| ***Household adversity pathway**** | | |
| Parental experience of domestic violence | 9 months<br>3 years | At 9 months and 3 years, main and partner respondent were asked if their partner ever used force in the relationship (yes, no). Domestic violence/abuse in the household was a binary variable defined as either partner reporting partner violence, at either 9 months or 3 years, regardless of family structure. |
| Parental substance misuse | 3 years | At 3 years, main and partner respondents were asked about their use of recreational drugs (Occasionally, |

| | | |
|---|---|---|
| | | regularly, never). A substance use problem in the household was defined as either main and partner respondent reporting regularly using recreational drugs at 3 years. |
| Parental mental illness | 3 years | |
| ***Parent-child pathway (continuous variables)*** | | |
| Frequency of parenting activities | 3 years | Main respondents were asked about whether they or someone else in the home do six types of activities with the child (reading, going to the library, learning the alphabet, drawing, learning numbers and learning songs), and the frequency of it. The frequency scores were converted into a continuous Home Learning Environment Index score (range: 0 to 42), using the procedure described in MCS data note 1 (11) which follows the definition of the Home Learning Environment (HLE) Index first described in Melhuish et al., (2008).<br><br>Main respondents were asked whether child goes to bed at regular times (Never, Sometimes, Usually Always). This was recoded to be in line with the HLE index which uses a 7-point score (1=Never/almost never , 3=Sometimes, 5=Usually, 7=Always)<br><br>The frequency scores for the HLE index and bedtime routine item were summed to provide the frequency of parenting activities (range: 0 to 49) |
| Parent-child relationship | 3 years | Parent-child relationship was measured at 3 years using the Child Parent Relationship Scale (Short form) which assesses relational conflict and closeness (13), 8 items assessing conflicts, 7 items assessing closeness on a 5-point Likert scale. Missing items due to "Can't say" or "Not applicable" was imputed to the midpoint score, if there were 3 or less "Can't say" or "Not applicable" responses. Each item was recoded so that (0=definitely does not apply and 4=definitely applies). Items on the conflicts scale were reverse coded, and then scores on 15 items summed to give the overall positive relationship (range: 0 to 60). Parent-child relationship was defined as the overall positive relationship reported by the main respondent, where a higher score indicates a more positive relationship. |
| Disciplinary parenting | 3 years | Main respondents were asked about frequency of using harsh parenting practices using 7 items from the Straus's Conflict Tactics Scale (1-never, rarely, once a month, at least once a week, 5-daily). Missing items due to "Can't say" or "Not applicable" was imputed to the midpoint score, if there were 2 or fewer responses in these categories. This included harsh parenting (shouting, smacking, telling off) and withdrawal tactics (ignoring, sending away to |

| | | |
|---|---|---|
| | | bedroom or naughty chair, taking away treats). Item scores were recoded to range between 0 and 4, and summed to create a continuous scale where higher scores indicate greater use of harsh parenting tactics (range: 0 to 28) |

* For the household adversity pathway, we do not include high frequency alcohol use as a mediating variable in this study as it shows a reverse social gradient in the Millennium Cohort Study. For the present analysis, this would imply an unrealistic intervention that increases the alcohol use in the low SES group to be equivalent to the high SES group. Alcohol is included as a confounding variable. Although previous work has also highlighted the importance of poverty and adversity, material deprivation is already captured in a separate pathway.

Although maternal attachment is available at 9 months, this is also captured by the closeness factor of the parent-child relationship.

**S3. Definitions of covariates in the Millennium Cohort Study (MCS)**

| Variable | Age assessed | |
|---|---|---|
| ***E-O or M-O confounders*** | | |
| Ethnicity | 9 months | UK Office for National Statistics five-category ethnic groups (White, Asian, Black, Mixed or Other) |
| Maternal mental health history | 9 months | Any longstanding illness falling in mental health or behavioural International Classification of Diseases (ICD-10) codes, or doctor's diagnosis of depression or severe anxiety (Yes, No) |
| Maternal childhood adversity | 9 months | Ever lived away from home in the care of local authority or foster carer (Yes, No) |
| Maternal age at cohort member's birth | 9 months | Years |
| ***M-O confounders*** | | |
| Language spoken at home | 9 months | English only or English and/or Other languages. |
| Number of children in household | 9 months | 1, 2, 3, or 4 or more children in household |
| ***Post-exposure M-O confounders or effect modifiers*** | | |
| Gestational age | 9 months | Weeks |
| Developmental delays | 9 months | Not meeting one or more motor, language or communicative milestone (out of 10 assessed) that 90 percent of the children in the MCS can do (Delay, No delays) |
| Child longstanding limiting illness | 3 years | Any reported long-term limiting illness (Yes, No) |
| Parental alcohol misuse | 9 months<br>3 years | High frequency alcohol use (Less than 5 times per week, 5-6 times per week or everyday), by main or partner respondent at either 9 months or 3 years (Yes, No) |
| ***Effect modifier*** | | |
| Child biological sex | 9 months | Male, Female |
| ***Auxiliary variables for imputation models*** | | |
| Voting status | 9 months<br>3 years | Ever not voted at the latest general election (Yes, No) |
| Breastfeeding | 9 months | Ever breastfed (Yes, No) |
| Household income | 3 years | OECD equivalised household income (Quintile 1 – lowest, to Quintile 5 – highest). |
| Housing tenure | 3 years | Owned, Rented/other |
| Accommodation type | 3 years | House/bungalow, Other |
| Occupational status | 3 years | Managerial/Professional, Intermediate, Small employers/self-employed, Lower supervisory/technical, Semi-routine/routine, Unemployed |
| Lone parenthood | 3 years | Yes, No |

**S4. Interventional disparity measures**

IDMs require the following assumptions with regards to the **mediator-outcome** relationship:

1) Consistency
2) No interference
3) No unmeasured confounding

The assumption of consistency in relation to the mediator outcome relationship states that within a group of children who share the same exposure, mediator and confounder levels, it we were to hypothetically intervene to shift the mediator distribution, it should produce the same mean outcome, as if those mediators were to naturally occur at the level we shifted it to. There are many ways we can hypothetically intervene on the mediators, and children live in the environments they do as a result of many possible reasons so it is difficult to conceive of a single intervention that would recreate the natural environment of children. Therefore, our intervention needs to be interpreted as a complex hypothetical intervention that randomly sets particular aspects of the childhood environment through many possible interventions. Despite the abstract intervention implied, effects are still relevant to policy, since we expect that many policies are conceivable, a range of different interventions are plausible in practice, and children and families may engage with interventions to varying extents and success.

The assumption of no interference means that the mental health outcome of one child is not affected by the early childhood environment of another child. This would occur if one child spends a lot of time in another child's environment, e.g. if they lived in the same household or were cared for by another child's parents. This is not the same as living in the same neighbourhood or attending the same childcare centre, where the same neighbourhood or childcare setting can represent each child's respective environment. We only included one child from each MCS family and excluded twins, and children are cared for primarily by parents, relatives or caregivers, and informal caregiving provided by one MCS family for another MCS child is unlikely. Interference may also occur if children who receive an early childhood intervention (i.e. improved aspect of their early childhood environment) achieve better outcomes, which in turn influence better outcomes in their peers who attend the same school or live in the same neighbourhood. Any potential interference due to improved outcomes of peers may lead to an underestimate of the true effect of intervening on the early childhood environment, because in reality, improving enough children's early childhood environment may impact outcomes of the community as a whole, similar to how vaccinating enough children would reduce the infection rate of non-vaccinated children.

In relation to the third assumption, no mediator-outcome confounding, we considered mediator-outcome confounders, including exposure-induced mediator-outcome confounders (i.e., intermediate confounders). However, residual confounding may be possible since maternal education may not fully capture the entirety of childhood SECs that confound the mediator outcome relationship (14).

***Additional notes on the use of IDMs in health inequalities research***

IDMs are suitable for research in health inequalities as the exposure (e.g. childhood socioeconomic circumstances) typically does not to fulfil the consistency assumption, where the exposure needs to be well-defined to inform a hypothetical intervention on how levels of the exposure would be changed. IDMs enables us to focus on unequal outcomes between different groups of people, which is highly relevant in the real-world, even without having a well-defined exposure on what is causing those inequalities (e.g. our exposure, unequal SECs, like socioeconomic position, is not well-defined as there are a myriad factors acting in complex ways to shape unequal outcomes).

IDMs may also be preferable in understanding pathways affecting socioeconomic inequality due to fewer assumptions in relation to confounding. Socioeconomic status has a pervasive effect on subsequent, downstream variables which can confound the mediator-outcome relationship (15). This is problematic when using natural effect models to understand the role of multiple mediators in mediating the effect of socioeconomic status on health, as it is not possible to adjust for an exposure-induced mediator-outcome confounder in estimation of natural direct and indirect effects. G-methods are able to adjust for an exposure-induced confounder, or the confounder may be included in the mediator vector (e.g. en bloc approach employed by (16)). However, these methods still rely on having no unmeasured mediator-mediator confounding. Secondly, unlike Interventional effects, IDMs do not require no unmeasured exposure-outcome or exposure-mediator confounding, because we are not interested in estimating the causal effect of the exposure on the outcome. In the estimation of IDMs, when some of the association of the exposure-outcome relationship might be due to residual confounding, the total adjusted association cannot be interpreted as the total causal effect. However, we are still able to address the question how much of the total adjusted association may be reduced by interventions on the mediating pathways, and compare these pathways (17).

Another mediation approach, controlled direct effects (CDEs), also require fewer assumptions on confounding. However, CDEs are not appropriate in the presence of exposure-mediator interactions - which is expected to be common in research on socioeconomic exposures. CDEs also cannot be used to calculate mediator specific indirect effects in the presence of mediator-mediator interactions, as the total resulting proportion can be greater than 1.

***IDMs in the context of multiple mediators***

A key strength of IDMs is in dealing with multiple mediators/mediating pathways. When some of the effects of a mediator are through a chain of mediators, the decomposition is such that the change in the AdjTA due to an intervention on any of the mediators in the chain is attributed to the most downstream mediator in the chain. For example, if household adversity affects mental health through parent-child relationships, the component of the adversity pathway would be captured in the parent-child relational pathway. Any association captured in the household adversity pathway would be associations due to any upstream influences on household adversity (e.g., home environment), but not due to another downstream mediator, (e.g, parent-child pathway). Importantly, this holds true even if underlying causal relationships differs from what is hypothesised because the estimation method is based on structural dependencies in the data, rather than how the order of mediators is specified. It must still be noted that the estimates only provide a descriptive decomposition of the exposure-outcome association explained via each mediator pathway, and do not prove causal relationships or ordering between the mediator pathways presented.

In dealing with mediator chains, IDMs also include a separate estimate of the mediated dependence component, when the effect of one mediator pathway depends on the presence of another pathway. In linear models, mediated dependence is non-zero when there are exposure-mediator and mediator-mediator interactions. Here, mediated dependence captures the nonlinear effects of multiple dimensions of risk or protective factors, but we do not further explore the dependencies between pathways.

**S5. Interventional Disparity Measures Definitions.**

**Intervention on early childhood mediators en bloc**

Define X childhood socioeconomic conditions (SECs), where x=0 (low), 1 (middle), 2 (high).

Define Y childhood mental health problems at 5 and 17 years (continuous) or define Y childhood psychiatric diagnosis at 5 and 17 years, where y=0 (absent), 1 (present).

Define $M = \{M_1 \dots M_n\}$ the vector of all the mediators of interest from the five pathways as

Define $C = \{C_1 \dots C_n\}$ the vector of covariates, which could include childhood SECs, parental mental health history, parental childhood adversity and maternal age, ethnicity, language spoken at home, number of children, parental high frequency alcohol use, preterm birth, early developmental delays, longstanding limiting illness or child biological sex.

Define Cx the exposure-specific vector of covariates including baseline confounders, and effect modifiers (sex, preterm birth, early developmental delays, longstanding limiting illness and parental high frequency alcohol use).

Define Cz the mediator-specific vector of covariates including baseline confounders, mediator-outcome confounders effect modifiers (sex, preterm birth, early developmental delays, longstanding limiting illness and parental high frequency alcohol use).

Define $M_c^x$ as a random draw from the **joint distribution of all the mediators**, amongst individuals where the exposure X=x, conditional on the pathway specific vector of covariates C, where C=c.

Define the interventional disparity measure indirect effect (IDM-IE, henceforth IE), the difference in expected childhood psychiatric diagnosis amongst the least advantaged (X=0), if the mediators were drawn from the joint distribution of those experienced by the most advantaged (X=2) $M_c^2$ versus if the mediators were drawn from the joint distribution of those experienced by the least advantaged (X=0) $M_c^0$, conditional on the exposure-specific vector of covariates C, where C=c.

$$IE = \sum_C \{E[Y(M_c^2)|X = 0, C = c] - E[Y(M_c^0)|X = 0, C = c]\}P(C = c)$$

Define the interventional disparity measure residual association (IDM-RA, henceforth RA), the difference in expected childhood psychiatric diagnosis between the least advantaged compared to the most advantaged that would remain, if the mediators were drawn from the joint distribution of mediators experienced by the most advantaged $M_c^2$.

$$RA = \sum_C \{E[Y(M_c^2)|X = 0, C = c] - E[Y(M_c^2)|X = 2, C = c]\}P(C = c)$$

Define the adjusted total association (AdjTA) the confounder adjusted association between the exposure and outcome, ie., the difference in mental health outcome between the least and most advantaged.

$$AdjTA = \sum_C \{E[Y(M_c^0)|X = 0, C = c] - E[Y(M_c^2)|X = 2, C = c]\}P(C = c)$$

The AdjTA, IE, and RA are related in this way:

$$AdjTA + IE = \text{RA}$$

**Decomposition via five pathways**

Further, define $Ma$ the two-category mediator class capturing neighbourhood pathway, where $Ma$ = Good, Poor conditions.

Define $Mb$ the mediator vector capturing the services and support pathway, where $Mb = \{Mb_1, Mb_2, Mb_3\}$, and $Mb_1$=attended antenatal classes (Yes, No), $Mb_2$=attended 6m centre-based childcare (Yes, No), and $Mb_3$=low social support (Yes, No)

Define $Mc$ the three-category mediator class capturing the material home environment pathway, where $Mc =$ Good, Overcrowded, Poor conditions

Define $Md$ the mediator capturing the adversity pathway, where $Md = \{Md_1, Md_2, Md_3\}$, and $Md_1$=parental experience of domestic violence (Yes, No) , $Md_2$=parental substance misuse (Yes, No), and $Md_3$=main respondent mental distress (continuous)

Define $Me$ the mediator capturing the material pathway, where $Me = \{Me_1, Me_2, Me_3\}$, and $Me_1$=frequency of parenting activities (continuous) , $Me_2$= disciplinary practices (continuous), and $Me_3$=positive parent-child relationship (continuous)

Define $Mz_C^x$ as a random draw from the marginal distribution of the mediator or mediator vector Mz, amongst individuals where the exposure X=x, conditional on the pathway specific vector of covariates C=c.

Define $IEz$ the pathway-specific indirect effect of the difference in outcome amongst the least advantaged (X=0) given exposure-specific vector of covariates C, where C=c, due to shifting $Mz$ from $Mz_C^0$ to $Mz_C^2$, but not any paths downstream of $Mz$ that involve any of the other mediator pathways.

$$IEa = \sum\nolimits_C \{E[Y(Ma_c^2, Mb_c^0, Mc_c^0, Md_c^0, Me_c^0)|X = \mathbf{0}, C = c] \\ - E[Y(Ma_c^0, Mb_c^0, Mc_c^0, Md_c^0, Me_c^0)|X = \mathbf{0}, C = c]\}P(C = c)$$

$$IEb = \sum\nolimits_C \{E[Y(Ma_c^0, Mb_c^2, Mc_c^0, Md_c^0, Me_c^0)|X = \mathbf{0}, C = c] \\ - E[Y(Ma_c^0, Mb_c^0, Mc_c^0, Md_c^0, Me_c^0)|X = \mathbf{0}, C = c]\}P(C = c)$$

$$IEc = \sum\nolimits_C \{E[Y(Ma_c^0, Mb_c^0, Mc_c^2, Md_c^0, Me_c^0)|X = \mathbf{0}, C = c] \\ - E[Y(Ma_c^0, Mb_c^0, Mc_c^0, Md_c^0, Me_c^0)|X = \mathbf{0}, C = c]\}P(C = c)$$

$$IEd = \sum\nolimits_C \{E[Y(Ma_c^0, Mb_c^0, Mc_c^0, Md_c^2, Me_c^0)|X = \mathbf{0}, C = c] \\ - E[Y(Ma_c^0, Mb_c^0, Mc_c^0, Md_c^0, Me_c^0)|X = \mathbf{0}, C = c]\}P(C = c)$$

$$IEe = \sum\nolimits_C \{E[Y(Ma_c^0, Mb_c^0, Mc_c^0, Md_c^0, Me_c^2)|X = \mathbf{0}, C = c] \\ - E[Y(Ma_c^0, Mb_c^0, Mc_c^0, Md_c^0, Me_c^0)|X = \mathbf{0}, C = c]\}P(C = c)$$

Define $MD$ the interventional disparity measure mediated dependence, which captures the difference in expected outcome amongst the least advantaged - due to the difference between the most and least advantaged in the conditional dependence between mediating pathways. In other words, the MD represents the difference in outcome amongst the least advantaged if the dependence between the mediator sets was what is seen in the most advantaged (first part of the expression), compared to if it was what is seen in the least advantaged.

$$MD = \sum\nolimits_C \{E[Y(M_c^2) - Y(Ma_c^2, Mb_c^2, Mc_c^2, Md_c^2, Me_c^2)|X = \mathbf{0}, C = C] \\ - E[Y(M_c^0) - Y(Ma_c^0, Mb_c^0, Mc_c^0, Md_c^0, Me_c^0)|X = \mathbf{0}, C = C]\}P(C = C)$$

The AdjTA can also be decomposed into the residual association (not due to any mediator pathway), the IE due to each pathway independently, as well as the mediated dependence between two or more pathways.

$$\mathrm{MD} = \mathrm{AdjTA} - RA - IEa - IEb - IEc - IEd - IEe$$

**S6. Estimation by Monte Carlo simulation with imputation of missing data**

I. Impute data.

II. Fit marginal and conditional mediator models, and outcome models using the original data

(1) First create models of each mediator. For the purpose of estimating each mediator within each pathway, $Mz_1$ is assumed to precede $Mz_2$, which precedes $Mz_3$. Fit models for each of the mediators in $Mz_n$, where z=a, b, c, d, e; n=1, 2… n, conditional on covariates Cz, and any of $Mz_j$ where j<n that precede $Mz_n$. This produces models of the mediators in $Mz$ which are marginal with respect to the other mediator pathways, i.e., $Mb, Mc, Md, Me$ are not adjusted for in models of $Ma_i$.

(2) Fit conditional models of each of the mediators $Mz_n$, where z = b, c, d, e; n=1, 2… n, conditional on $X$, $Cz$, $Ma$ and all mediators in any preceding pathway. The conditional model for $Ma$ is the same as the marginal model, since there are no preceding pathways.

(3) Fit models of the outcome conditional on all the mediators, baseline confounders and effect modifiers.

III. Create the simulation dataset with only individuals with X=0 and X=2, so that that marginalisation over covariates is performed based on the covariates observed in individuals with X=0 and X=2 only.

IV. Expand data by N-fold, so that mediator draws and outcome prediction are simulated N times for each individual.

V. Simulate mediator draws based on the marginal mediator distribution

(1) Using marginal models of each mediator $Mz_n$ where z = a, b, c, d, e; n=1, 2… n, draw for each subject *i* given $C = Ci$, for $X = 0$ and $X$ =2, on the expanded dataset to give $Mz_c^0$ and $Mz_c^2$.

VI. Simulate mediator draws based on the joint mediator distribution using recursive mediator draws to simulate draws of mediator $Mz$ dependent on draws of mediators in an earlier pathway.

(2) Using conditional models of each mediator $Mb_n$, n=1, 2… n, draw for each subject *i* given $C = Ci$ and $Ma = Ma_c^x$, for $X = 0$ and $X$ =2, on the expanded dataset to give $Mz'^0_c$ and $Mz'^2_c$.

(3) Repeat for $Mc_n$ conditional on $C = Ci, Ma$ and $Mb'^x_c$ to obtain $Mc'^0_c$ and $Mc'^2_c$.

(4) Repeat for $Md_n$ conditional on $C = Ci, Ma$, $Mb'^x_c$, $Mc'^x_c$.to obtain $Md'^0_c$ and $Md'^2_c$.

(5) Repeat for $Me_n$ conditional on $C = Ci, Ma$, $Mb'^x_c$, $Mc'^x_c$, $Md'^x_c$ to obtain $Me'^0_c$ and $Me'^2_c$.

(6) The joint distribution of mediators $M_c^x$ when $X = 0$ and $X$ =2 is given by $M_c^0 = \{Ma_c^0, Mb'^0_c, Mc'^0_c, Md'^0_c, Me'^0_c\}$ and $M_c^1 = \{Ma_c^1, Mb'^1_c, Mc'^1_c, Md'^1_c, Me'^1_c\}$

*Note: This is based on applying the probability chain rule recursively for five variables, using independent draws of* $Ma$ $(Ma_c^x)$ *and recursive draws of* $Mb$ $(Mb'^x_c)$, $Mc$ $(Mc'^x_c)$, $Md$ $(Md'^x_c)$ *and* $Me$ $(Me'^x_c)$ *conditional on any preceding mediators.*

$$
\begin{aligned}
P(E,D,C,B,A) &= P(E|A,B,C,D)P(A,B,C,D) = P(E|A,B,C,D)P(D|A,B,C)P(A,B,C) \\
&= P(E|A,B,C,D)P(D|A,B,C)P(C|A,B)P(A,B) \\
&= P(E|A,B,C,D)P(D|A,B,C)P(C|A,B)P(B|A)P(A)
\end{aligned}
$$

VII. Predict outcomes in the N-fold expanded data based on terms required in equations.

(1) *AdjTA*. Predict the conditional outcomes given each subject's $C = Ci$, for $M = M_c^0, X = 0$, and $M = M_c^2, X = 2$.

(2) *IE*. Predict the conditional outcomes given each subject's $C = Ci$, for $M = M_c^2, X = 0$, and $M = M_c^0, X = 0$.

(3) *RA*. Predict the conditional outcomes given each subject's $C = Ci$, for $M = M_c^2, X = 0$, and $M = M_c^2, X = 2$.

(4) *IEa.* Predict the conditional outcomes given each subject's $C = Ci$, for $M = \{Ma_c^2, Mb_c^0, Mc_c^0, Md_c^0, Me_c^0\}, X = 0$, and $M = \{Ma_c^0, Mb_c^0, Mc_c^0, Md_c^0, Me_c^0\}, X = 0$.

(5) *IEb*. Predict the conditional outcomes given each subject's $C = Ci$, for $M = \{Ma_c^0, Mb_c^2, Mc_c^0, Md_c^0, Me_c^0\}, X = 0$, and $M = \{Ma_c^0, Mb_c^0, Mc_c^0, Md_c^0, Me_c^0\}, X = 0$.
(6) *IEc*. Predict the conditional outcomes given each subject's $C = Ci$, for $M = \{Ma_c^0, Mb_c^0, Mc_c^2, Md_c^0, Me_c^0\}, X = 0$, and $M = \{Ma_c^0, Mb_c^0, Mc_c^0, Md_c^0, Me_c^0\}, X = 0$.
(7) *IEd*. Predict the conditional outcomes given each subject's $C = Ci$, for $M = \{Ma_c^0, Mb_c^0, Mc_c^0, Md_c^2, Me_c^0\}, X = 0$, and $M = \{Ma_c^0, Mb_c^0, Mc_c^0, Md_c^0, Me_c^0\}, X = 0$.
(8) *IEe*. Predict the conditional outcomes given each subject's $C = Ci$, for $M = \{Ma_c^0, Mb_c^0, Mc_c^0, Md_c^0, Me_c^2\}, X = 0$, and $M = \{Ma_c^0, Mb_c^0, Mc_c^0, Md_c^0, Me_c^0\}, X = 0$
(9) *MD*. Predict the conditional outcomes given each subject's $C = Ci$, for $M = M_c^2, X = 0$; $M = \{Ma_c^2, Mb_c^2, Mc_c^2, Md_c^2, Me_c^2\}, X = 0$; $M = M_c^0, X = 0$ and $\{Ma_c^0, Mb_c^0, Mc_c^0, Md_c^0, Me_c^0\}, X = 0$.

VIII. Calculate Interventional Disparity Measures according to equations by obtaining conditional mean difference in outcomes, performing marginalisation over covariates

(1) According to the equations for *AdjTA*, IE, RA, IEz and MD, obtain the difference in the outcomes for each subject.
(2) Obtain the conditional mean difference in outcomes by averaging over all subjects in the expanded dataset. This performs marginalisation over C=Ci, for individuals with observed X=0 and X=2.
(3) Optional check: obtain the sum of IEz and MD, to check that it gives the same value as the IE.

IX. Repeat Steps I to VIII 500 times, including a bootstrap of the imputed data in Step I by randomly selecting with replacement. IDMs are therefore simulated using the bootstrap replicate of the original data in subsequent steps.

X. Obtain confidence intervals of Monte Carlo simulation of IDMs.

(1) Check if IDMs are normally distributed across the bootstrapped replicates.
(2) If so, calculate the bias-corrected normal approximation of confidence intervals using the norm.ci function from the *boot* package.
(3) If not, perform another 500 bootstraps and obtain the bias-corrected percentile confidence intervals for 1000 bootstrapped replicates, using the *perc.ci* function from the *boot* package (see note)

*Note: Bias may arise because the bootstrap distribution is based on resampling the observed data, which may not perfectly represent the population. The bootstrap bias estimate reflects the expected difference between the estimator computed in the sample and the population parameter. Instead of centring the 95% confidence interval symmetrically around the observed estimate, bias-corrected percentile confidence intervals are constructed by adjusting the 2.5 and 97.5 percentile cut points of the bootstrap distribution to account for the estimated bias, rather than by centring the interval symmetrically around the observed estimate. The resulting bias-corrected 95% confidence interval represents a range with approximately 95% coverage for the true population value under the bootstrap assumptions. The estimate from the observed sample remains the point estimate of the population parameter; the mean of the bootstrap estimates is not used as the point estimate, as it estimates the expected value of the estimator rather than the parameter itself.*

Example:

Models of the mediators were used to simulate the distribution of each mediator. We then obtained mediator values based on draws from the joint distribution of all mediators, given an exposure and covariate status. Then the outcome model was used to predict the mental health outcome based on a specified exposure value, the simulated mediators, and the individual's original covariate status. Repeating this for the two exposure values (High and Low SEC), and mediator draws based on different exposure values gives the terms required to calculate each IDM in equations in Appendix S5.

In the calculation of the IE, we simulate the mental health outcome in Low SEC individuals by drawing 200 random mediator values from the High SEC distribution, given the individual's original covariate status, and repeated this 200 times. Then the outcome model was used to predict the mental health outcome in the counterfactual scenario that all individuals have Low SEC exposure, randomly drawn High SEC mediators, given their original covariate status. We repeat mediator draws and expected outcome in the scenario that all individuals have Low SEC exposure, Low SEC mediators, given their original covariate status. The IDM Indirect Effect is calculated as the expected difference in the outcomes in the scenario that individuals have Low

SEC exposure, High SEC mediators, given versus if individuals have Low SEC exposure, Low SEC mediators, given their original covariate status.

Similarly, we repeated this for the two exposure values (High and Low SEC), and mediator draws based on different exposure values to obtain the terms for calculating pathway-specific IDMs in equations in Appendix S5.

**S7. References**


1. Goodman R, Ford T, Simmons H, Gatward R, Meltzer H. Using the Strengths and Difficulties Questionnaire (SDQ) to screen for child psychiatric disorders in a community sample. The British Journal of Psychiatry. 2000 Dec;177(6):534–9.
2. Kessler RC, Andrews G, Colpe LJ, Hiripi E, Mroczek DK, Normand SLT, et al. Short screening scales to monitor population prevalences and trends in non-specific psychological distress. Psychol Med. 2002 Aug;32(6):959–76.
3. Kessler RC, Barker PR, Colpe LJ, Epstein JF, Gfroerer JC, Hiripi E, et al. Screening for Serious Mental Illness in the General Population. Archives of General Psychiatry. 2003 Feb 1;60(2):184–9.
4. Green JG, Gruber MJ, Sampson NA, Zaslavsky AM, Kessler RC. Improving the K6 short scale to predict serious emotional disturbance in adolescents in the USA. Int J Methods Psychiatr Res. 2010 June;19 Suppl 1(Suppl 1):23–35.
5. Mewton L, Kessler RC, Slade T, Hobbs MJ, Brownhill L, Birrell L, et al. The psychometric properties of the Kessler Psychological Distress Scale (K6) in a general population sample of adolescents. Psychological Assessment. 2016;28(10):1232–42.
6. Nussbaum M. Poverty and Human Functioning: Capabilities as Fundamental Entitlements. In: Poverty and Inequality [Internet]. Stanford University Press; 2006. Available from: https://chicagounbound.uchicago.edu/book_chapters/796/
7. Rod NH, Broadbent A, Rod MH, Russo F, Arah OA, Stronks K. Complexity in Epidemiology and Public Health. Addressing Complex Health Problems Through a Mix of Epidemiologic Methods and Data. Epidemiology. 2023 July;34(4):505.
8. Green MJ, Pearce A, Parkes A, Robertson E, Katikireddi SV. Pre-school childcare and inequalities in child development. SSM - Population Health. 2021 June 1;14:100776.
9. Department for Work and Pensions. Family Resources Survey: background information and methodology [Internet]. 2024 [cited 2024 Nov 4]. Available from: https://www.gov.uk/government/statistics/family-resources-survey-financial-year-2022-to-2023/family-resources-survey-background-information-and-methodology
10. McKay S. Review of the child material deprivation items in the family resources survey: a report of research carried out by Birmingham University on behalf of the Department for Work and Pensions [Internet]. London: Department for Work and Pensions; 2011 [cited 2024 Nov 4]. Available from: http://dera.ioe.ac.uk/3644/1/rrep746.pdf
11. Rochebrochard E. Millennium Cohort Study Data Note 1: The home learning environment as measured at age 3. London: Centre for Longitudinal Studies; 2012 July.
12. Melhuish EC, Phan MB, Sylva K, Sammons P, Siraj-Blatchford I, Taggart B. Effects of the Home Learning Environment and Preschool Center Experience upon Literacy and Numeracy Development in Early Primary School. Journal of Social Issues. 2008;64(1):95–114.
13. Driscoll K, Pianta RC. Mothers' and fathers' perceptions of conflict and closeness in parent-child relationships during early childhood. Journal of Early Childhood and Infant Psychology. 2011;7:1–24.
14. Galobardes B, Shaw M, Lawlor DA, Lynch JW, Smith GD. Indicators of socioeconomic position (part 1). Journal of Epidemiology & Community Health. 2006 Jan 1;60(1):7–12.
15. Naimi AI, Schnitzer ME, Moodie EEM, Bodnar LM. Mediation Analysis for Health Disparities Research. American Journal of Epidemiology. 2016 Aug 15;184(4):315–24.
16. Straatmann VS, Lai E, Lange T, Campbell MC, Wickham S, Andersen AMN, et al. How do early-life factors explain social inequalities in adolescent mental health? Findings from the UK Millennium Cohort Study. J Epidemiol Community Health. 2019 Nov 1;73(11):1049–60.
17. Micali N, Daniel RM, Ploubidis GB, De Stavola BL. Maternal Prepregnancy Weight Status and Adolescent Eating Disorder Behaviors: A Longitudinal Study of Risk Pathways. Epidemiology. 2018 July;29(4):579–89.

**S8. Supplemental Figures and Tables**

**Figure S1. Neighbourhood (top) and material (bottom) class item probabilities**

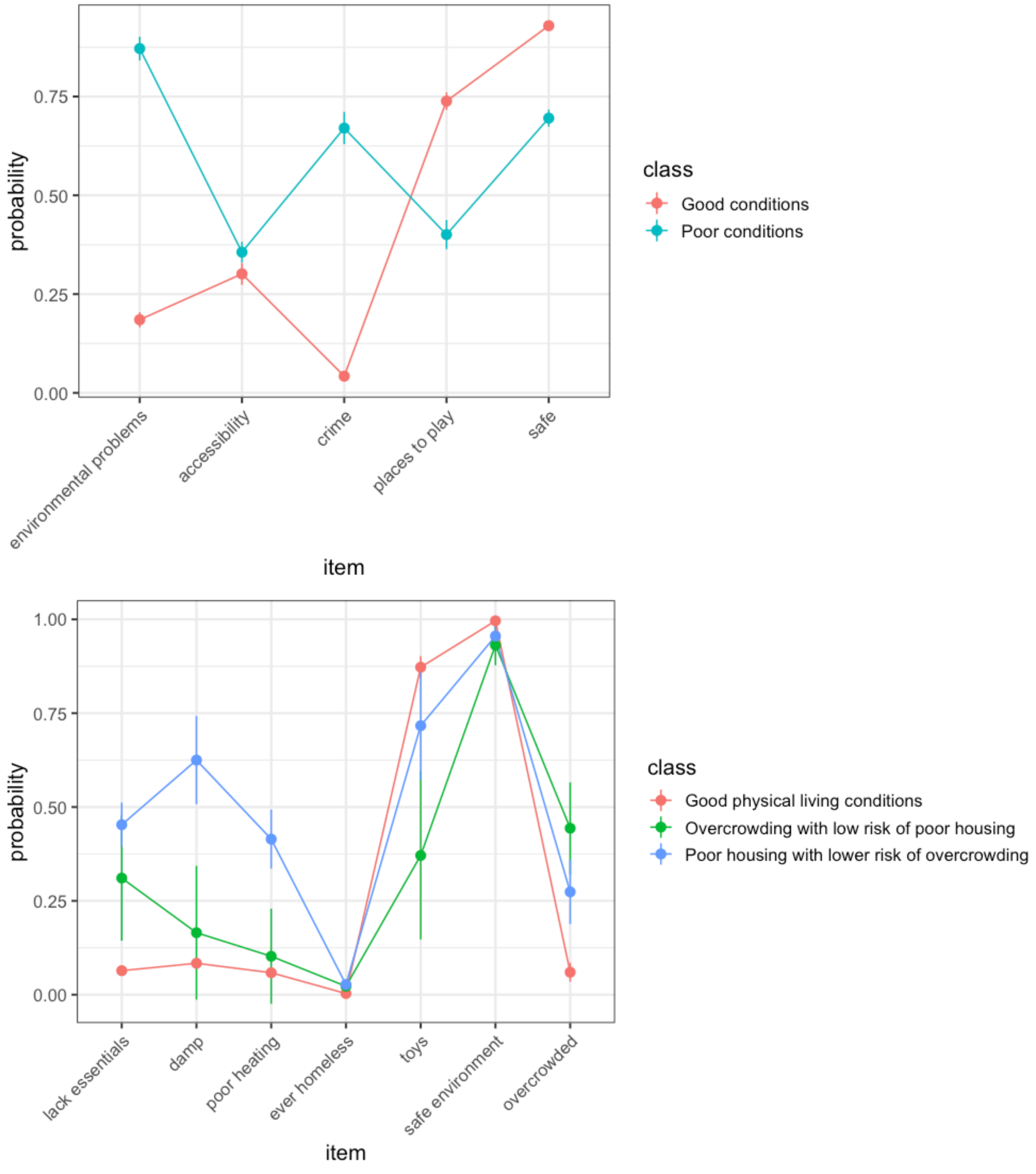

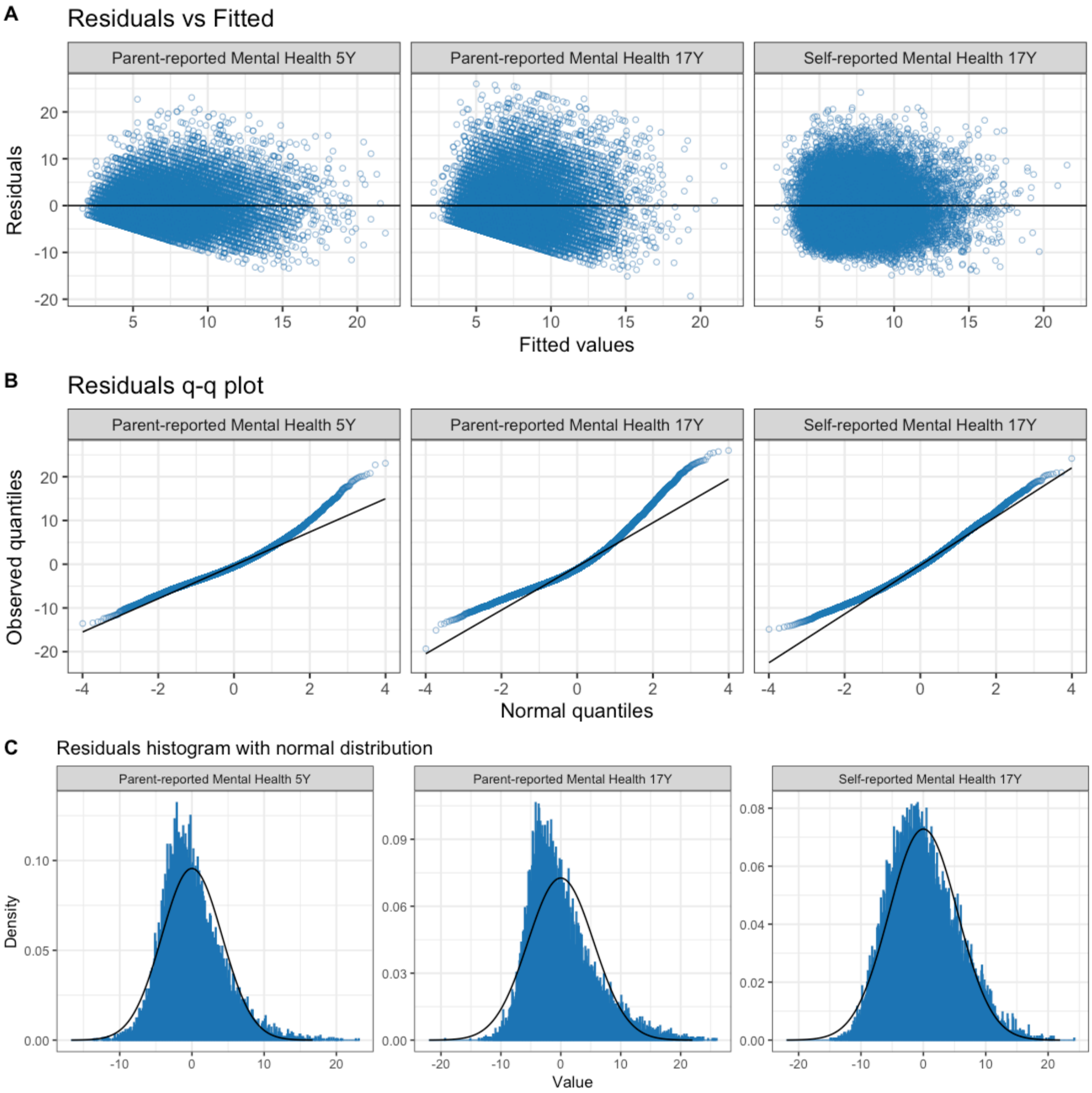


**Figure S2. Residual plots for outcome models – original scale. Approximately normal and randomly spread, but residual q-q plot indicates positive skew in the residuals.**

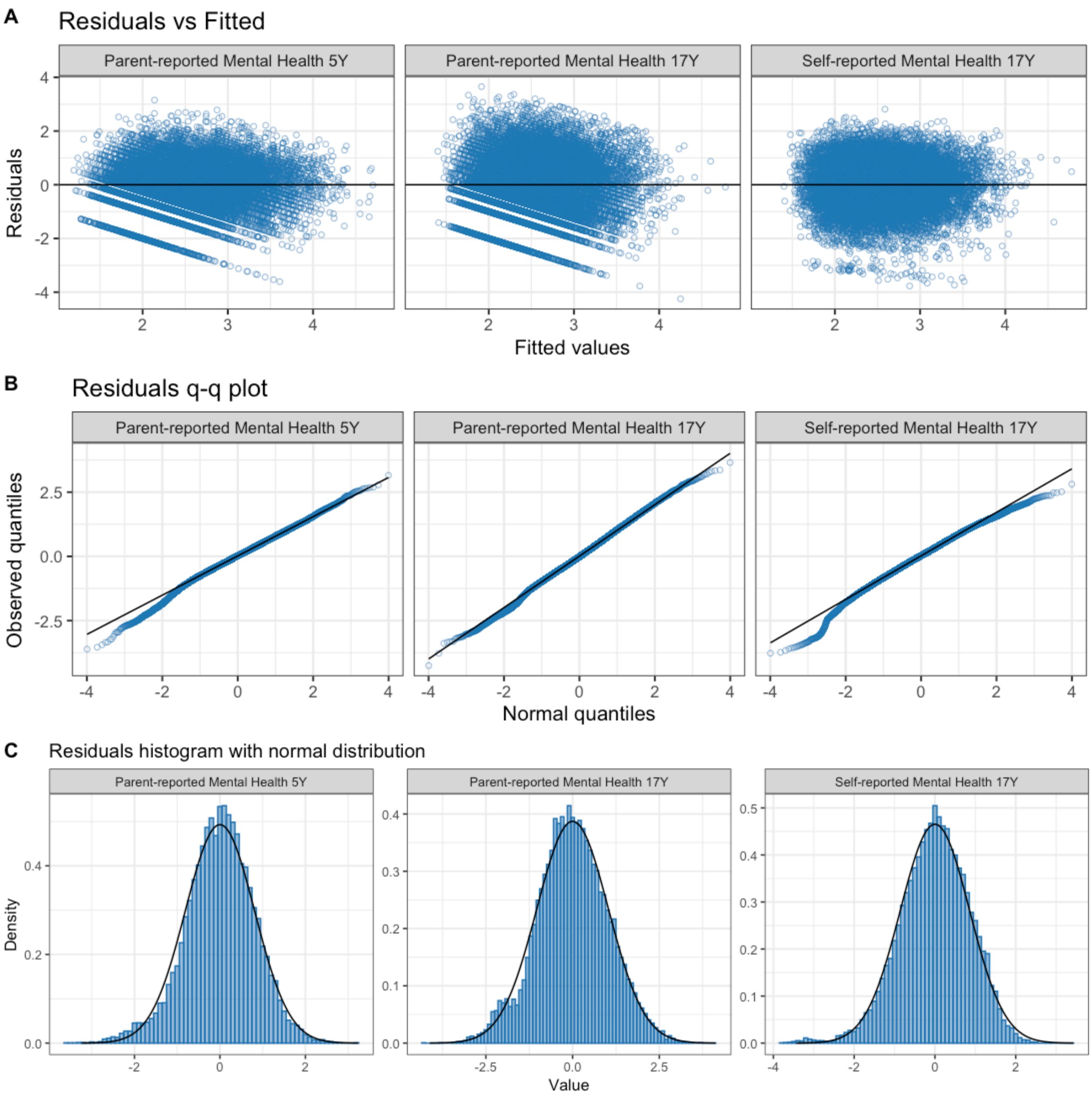


**Figure S3. Residual plots for outcome models with square-root transformation**

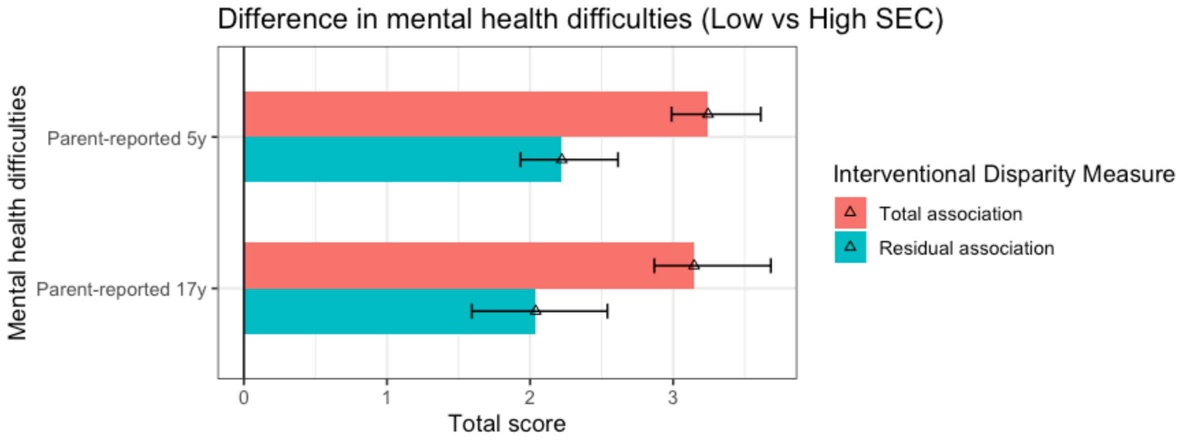


**Figure S4. IDM Total association and residual association after intervening *en bloc* on early childhood pathways**

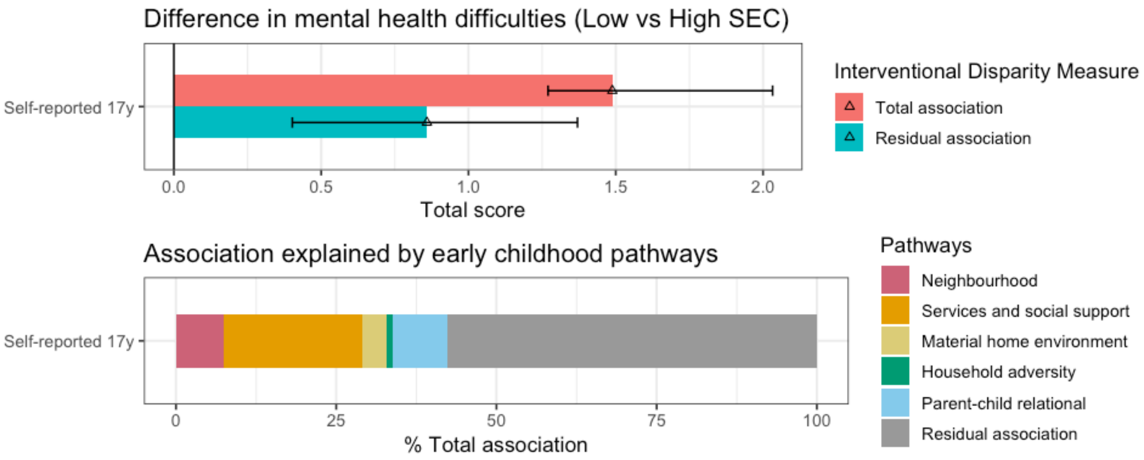


**Figure S5. Self-reported 17y mental health difficulties- IDM Total association, residual association (Top) and decomposition of IE via five pathways (bottom)**

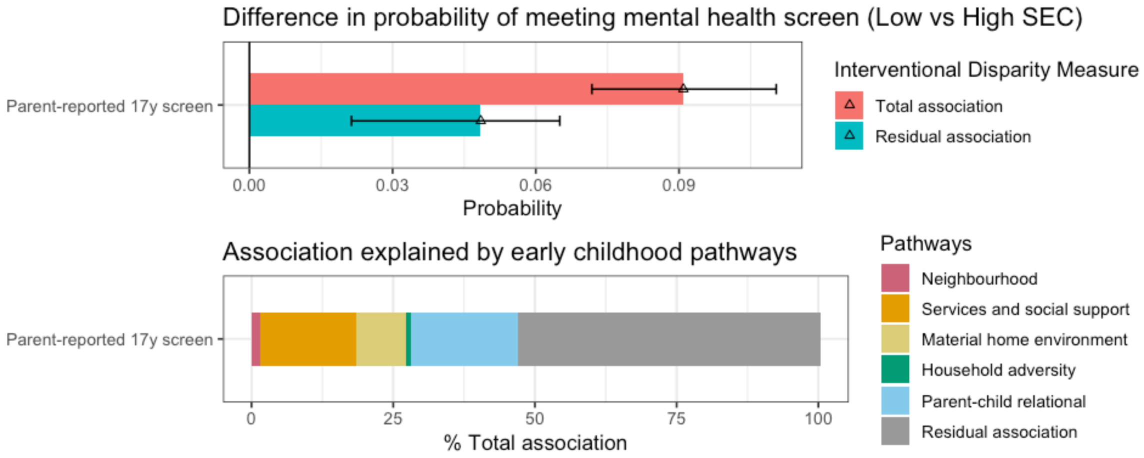


**Figure S6. Parent-reported 17y screen - IDM Total association, residual association (Top) and decomposition of IE via five pathways (bottom)**

**Table S1. Latent Class Analysis model fit indices for up to 3 classes**

| Class | Entropy | AIC | BIC | LL | minus2LL | Observations | Smallest class |
|---|---|---|---|---|---|---|---|
| **Neighbourhood pathway** | | | | | | | |
| 1 | 2.909526 | 84601.55 | 84639.47 | -42295.8 | 84591.55 | 14537 | NA |
| 2 | 2.768179 | 80478.35 | 80561.78 | -40228.2 | 80456.35 | 14537 | 3464 |
| 3 | Non-convergence | | | | | | |
| **Material pathway** | | | | | | | |
| 1 | 2.297968 | 61208.89 | 61261.37 | -30597.4 | 61194.89 | 13315 | NA |
| 2 | 2.234987 | 59519.3 | 59631.75 | -29744.7 | 59489.3 | 13315 | 2860 |
| 3 | 2.226323 | 59312.08 | 59484.5 | -29633 | 59266.08 | 13315 | 1072 |

**Table S2. Simulation of Monte Carlo error using complete case samples. We selected 200 expansions as this reduced Monte Carlo error to 2 decimal places and 0 decimal places for percentages, allowing for rounding error. Error in the MD, since the MD was small and the percentage IE did not change substantially. We did not further increase the simulations due to the large computational cost. We report estimates based on the error in the selected number of decimal places.**

| Y | expansions | seed | IE | RA | AdjTA | IEa | IEb | IEc | IEd | IEe | MD |
|---|---|---|---|---|---|---|---|---|---|---|---|
| Y17 | 100 | 1 | -0.9354 | 1.6501 | 2.5855 | -0.1547 | -0.4222 | -0.1279 | -0.0071 | -0.2165 | -0.0070 |
| Y17 | 100 | 2 | -0.9405 | 1.6493 | 2.5898 | -0.1537 | -0.4230 | -0.1291 | -0.0089 | -0.2092 | -0.0167 |
| Y17 | 100 | 3 | -0.9282 | 1.6455 | 2.5737 | -0.1534 | -0.4213 | -0.1275 | -0.0061 | -0.2161 | -0.0037 |
| Y5 | 100 | 1 | -0.8163 | 1.8734 | 2.6897 | -0.1439 | -0.0890 | -0.0639 | -0.0448 | -0.5417 | 0.0671 |
| Y5 | 100 | 2 | -0.8328 | 1.8705 | 2.7033 | -0.1431 | -0.0918 | -0.0643 | -0.0426 | -0.5375 | 0.0465 |
| Y5 | 100 | 3 | -0.8179 | 1.8722 | 2.6901 | -0.1433 | -0.0907 | -0.0643 | -0.0427 | -0.5424 | 0.0656 |
| Y17 | 200 | 1 | -0.9368 | 1.6467 | 2.5834 | -0.1532 | -0.4236 | -0.1273 | -0.0077 | -0.2153 | -0.0097 |
| Y17 | 200 | 2 | -0.9440 | 1.6449 | 2.5890 | -0.1534 | -0.4229 | -0.1272 | -0.0077 | -0.2090 | -0.0237 |
| Y17 | 200 | 3 | -0.9387 | 1.6461 | 2.5848 | -0.1540 | -0.4253 | -0.1299 | -0.0056 | -0.2141 | -0.0098 |
| Y5 | 200 | 1 | -0.8227 | 1.8718 | 2.6945 | -0.1430 | -0.0896 | -0.0650 | -0.0420 | -0.5435 | 0.0604 |
| Y5 | 200 | 2 | -0.8303 | 1.8715 | 2.7018 | -0.1435 | -0.0904 | -0.0645 | -0.0431 | -0.5384 | 0.0497 |
| Y5 | 200 | 3 | -0.8253 | 1.8724 | 2.6977 | -0.1450 | -0.0904 | -0.0655 | -0.0418 | -0.5421 | 0.0595 |

| Y | expansions | seed | IE | RA | AdjTA | IEa (% AdjTA) | IEb (% AdjTA) | IEc (% AdjTA) | IEd (% AdjTA) | IEe (% AdjTA) | MD (% AdjTA) |
|---|---|---|---|---|---|---|---|---|---|---|---|
| Y17 | 100 | 1 | -36.18 | 63.82 | 100.00 | -5.98 | -16.33 | -4.95 | -0.27 | -8.37 | -0.27 |
| Y17 | 100 | 2 | -36.32 | 63.68 | 100.00 | -5.93 | -16.33 | -4.98 | -0.34 | -8.08 | -0.64 |
| Y17 | 100 | 3 | -36.06 | 63.94 | 100.00 | -5.96 | -16.37 | -4.96 | -0.24 | -8.40 | -0.15 |
| Y5 | 100 | 1 | -30.35 | 69.65 | 100.00 | -5.35 | -3.31 | -2.37 | -1.67 | -20.14 | 2.49 |
| Y5 | 100 | 2 | -30.81 | 69.19 | 100.00 | -5.29 | -3.40 | -2.38 | -1.58 | -19.88 | 1.72 |
| Y5 | 100 | 3 | -30.40 | 69.60 | 100.00 | -5.33 | -3.37 | -2.39 | -1.59 | -20.16 | 2.44 |
| Y17 | 200 | 1 | -36.26 | 63.74 | 100.00 | -5.93 | -16.40 | -4.93 | -0.30 | -8.34 | -0.37 |
| Y17 | 200 | 2 | -36.46 | 63.54 | 100.00 | -5.93 | -16.34 | -4.91 | -0.30 | -8.07 | -0.92 |
| Y17 | 200 | 3 | -36.32 | 63.68 | 100.00 | -5.96 | -16.45 | -5.03 | -0.22 | -8.28 | -0.38 |
| Y5 | 200 | 1 | -30.53 | 69.47 | 100.00 | -5.31 | -3.33 | -2.41 | -1.56 | -20.17 | 2.24 |
| Y5 | 200 | 2 | -30.73 | 69.27 | 100.00 | -5.31 | -3.35 | -2.39 | -1.59 | -19.93 | 1.84 |
| Y5 | 200 | 3 | -30.59 | 69.41 | 100.00 | -5.37 | -3.35 | -2.43 | -1.55 | -20.09 | 2.21 |

**Table S3. Number and percentage of individuals with 1 to 6 available mental health outcomes available from 5 to 17 years (parent-reported SDQ at 5, 7, 11, 14, and 17 years, and self-reported SDQ at 17 years)**

| Number of mental health measures available | n | % |
|---|---|---|
| 1 | 1321 | 8.5 |
| 2 | 1622 | 10.5 |
| 3 | 1826 | 11.8 |
| 4 | 2138 | 13.8 |
| 5 | 2103 | 13.6 |
| 6 | 6503 | 41.9 |

**Table S4. Missing variables imputed using multiple imputation**

| Variable | n | % |
|---|---|---|
| ***Exposure*** | | |
| education_9m | 0 | 0.0 |
| ***Covariates*** | | |
| PTTYPE2 | 0 | 0.0 |
| sex | 0 | 0.0 |
| ethnicity5 | 803 | 5.2 |
| maternal_age | 2 | 0.0 |
| maternal_mh_history | 0 | 0.0 |
| care_experience | 0 | 0.0 |
| language | 1 | 0.0 |
| children_in_household | 1643 | 10.6 |
| alc | 0 | 0.0 |
| gestation | 538 | 3.5 |
| lli | 1727 | 11.1 |
| delay | 378 | 2.4 |
| ***Mediators*** | | |
| nb_class | 0 | 0.0 |
| mat_class | 0 | 0.0 |
| ei_ant | 908 | 5.9 |
| ei_cc | 1734 | 11.2 |
| ei_soc | 596 | 3.8 |
| dv | 2329 | 15.0 |
| subs | 2532 | 16.3 |
| mh_main | 3157 | 20.4 |
| em_dis | 4398 | 28.4 |
| em_rel | 3114 | 20.1 |
| em_par | 1737 | 11.2 |
| ***Mental health outcomes*** | | |
| Tot_3 | 1493 | 9.6 |
| Tot_4 | 2671 | 17.2 |
| Tot_5 | 3267 | 21.1 |
| Tot_6 | 4681 | 30.2 |
| Tot_7 | 6640 | 42.8 |
| cm_sdqc | 6198 | 40.0 |
| ***Auxilliary variables*** | | |
| ever_not_voted | 1085 | 7.0 |
| accom_type | 378 | 2.4 |
| housing_tenure | 2 | 0.0 |
| breastfed | 22 | 0.1 |
| occupation_status | 756 | 4.9 |
| family_structure | 1 | 0.0 |
| income_9m | 28 | 0.2 |

**Table S5. Descriptive statistics of multiple imputation and non-weighted complete case samples**

| Characteristic | Main analytic sample N = 15,513[1] | 5y complete case N = 8,477[1] | 17y complete case N = 5,612[1] |
|---|---|---|---|
| sec | | | |
| High | 2,668 (17.2%) | 1,855 (21.9%) | 1,428 (25.4%) |
| Middle | 10,037 (64.7%) | 5,833 (68.8%) | 3,730 (66.5%) |
| Low | 2,808 (18.1%) | 789 (9.3%) | 454 (8.1%) |
| education_9m | | | |
| Degree plus | 2,668 (17.2%) | 1,855 (21.9%) | 1,428 (25.4%) |
| Diploma | 1,394 (9.0%) | 951 (11.2%) | 654 (11.7%) |
| A level | 1,537 (9.9%) | 990 (11.7%) | 680 (12.1%) |
| GCSE A-C | 5,404 (34.8%) | 3,092 (36.5%) | 1,927 (34.3%) |
| GCSE D-G | 1,702 (11.0%) | 800 (9.4%) | 469 (8.4%) |
| No qualifications | 2,808 (18.1%) | 789 (9.3%) | 454 (8.1%) |
| income_9m | | | |
| 1 | 3,656 (23.6%) | 910 (10.7%) | 524 (9.3%) |
| 2 | 3,392 (21.9%) | 1,660 (19.6%) | 1,021 (18.2%) |
| 3 | 2,966 (19.2%) | 1,881 (22.2%) | 1,218 (21.7%) |
| 4 | 2,842 (18.4%) | 2,056 (24.3%) | 1,413 (25.2%) |
| 5 | 2,629 (17.0%) | 1,964 (23.2%) | 1,433 (25.5%) |
| poverty_9m | | | |
| Above 60% median | 10,055 (64.9%) | 6,776 (80.0%) | 4,578 (81.6%) |
| Below 60% median | 5,430 (35.1%) | 1,695 (20.0%) | 1,031 (18.4%) |
| housing_tenure | | | |
| Owned | 9,642 (62.2%) | 6,548 (77.2%) | 4,478 (79.8%) |
| Rented/other | 5,869 (37.8%) | 1,929 (22.8%) | 1,134 (20.2%) |
| occupation_status | | | |
| Managerial/Professional | 5,373 (36.4%) | 4,005 (48.0%) | 2,895 (52.3%) |
| Intermediate | 1,451 (9.8%) | 958 (11.5%) | 608 (11.0%) |
| Small employers/self-employed | 1,013 (6.9%) | 630 (7.5%) | 414 (7.5%) |
| Lower supervisory/technical | 1,101 (7.5%) | 691 (8.3%) | 413 (7.5%) |
| Semi-routine/routine | 2,570 (17.4%) | 1,302 (15.6%) | 790 (14.3%) |
| Unemployed | 3,249 (22.0%) | 759 (9.1%) | 416 (7.5%) |
| lone_parent | | | |
| Natural/reconstituted family | 12,716 (82.0%) | 8,043 (94.9%) | 5,363 (95.6%) |
| Lone parent | 2,797 (18.0%) | 434 (5.1%) | 249 (4.4%) |
| sex | | | |
| Female | 7,560 (48.7%) | 4,146 (48.9%) | 2,811 (50.1%) |
| Male | 7,953 (51.3%) | 4,331 (51.1%) | 2,801 (49.9%) |
| ethnicity5 | | | |
| White | 13,255 (90.1%) | 8,111 (95.7%) | 5,327 (94.9%) |
| Asian | 166 (1.1%) | 41 (0.5%) | 27 (0.5%) |
| Black | 366 (2.5%) | 132 (1.6%) | 98 (1.7%) |
| Mixed | 653 (4.4%) | 153 (1.8%) | 130 (2.3%) |
| Other | 270 (1.8%) | 40 (0.5%) | 30 (0.5%) |
| maternal_teen_firstchild | 1,086 (7.0%) | 361 (4.3%) | 184 (3.3%) |
| maternal_age | 28.4 (5.9) | 29.4 (5.4) | 29.8 (5.3) |
| maternal_mh_history | 3,927 (25.3%) | 2,045 (24.1%) | 1,278 (22.8%) |
| care_experience | 247 (1.6%) | 90 (1.1%) | 46 (0.8%) |
| children_in_household | | | |
| 1 | 3,512 (25.3%) | 1,998 (23.6%) | 1,287 (22.9%) |
| 2 | 6,245 (45.0%) | 4,178 (49.3%) | 2,826 (50.4%) |
| 3 | 2,670 (19.3%) | 1,616 (19.1%) | 1,071 (19.1%) |

| | | | |
|---|---|---|---|
| 4 or more | 1,443 (10.4%) | 685 (8.1%) | 428 (7.6%) |
| gestation | 39.4 (2.0) | 39.5 (1.9) | 39.5 (1.9) |
| preterm | | | |
| Preterm | 1,672 (11.0%) | 739 (8.7%) | 493 (8.8%) |
| Term | 13,463 (89.0%) | 7,738 (91.3%) | 5,119 (91.2%) |
| language | | | |
| English only | 13,344 (86.0%) | 7,919 (93.4%) | 5,196 (92.6%) |
| English and other languages | 2,168 (14.0%) | 558 (6.6%) | 416 (7.4%) |
| lli | 421 (3.1%) | 224 (2.6%) | 155 (2.8%) |
| delay | 2,343 (15.5%) | 1,249 (14.7%) | 833 (14.8%) |

**Table S6. Survey-weighted mediator distribution by exposure status 5y complete case sample (Unweighted n=8477)**

| Characteristic | High, N = 1,992[1] | Middle, N = 6,319[1] | Low, N = 777[1] | p-value[2] |
|---|---|---|---|---|
| **Neighbourhood pathway** | | | | |
| Neighbourhood class | | | | <0.001 |
| Good conditions | 1,760 (88.4%) | 5,091 (80.6%) | 519 (66.8%) | |
| Poor conditions | 231 (11.6%) | 1,228 (19.4%) | 258 (33.2%) | |
| **Services and social support pathway** | | | | |
| Attended antenatal classes | | | | <0.001 |
| No | 870 (43.7%) | 3,861 (61.1%) | 623 (80.2%) | |
| Yes | 1,122 (56.3%) | 2,457 (38.9%) | 154 (19.8%) | |
| Attended 6 months centre-based childcare by 32 months | | | | <0.001 |
| No | 1,253 (62.9%) | 5,223 (82.7%) | 728 (93.6%) | |
| Yes | 739 (37.1%) | 1,096 (17.3%) | 49 (6.4%) | |
| Low social support | | | | <0.001 |
| No | 1,849 (92.8%) | 6,090 (96.4%) | 744 (95.7%) | |
| Yes | 143 (7.2%) | 229 (3.6%) | 33 (4.3%) | |
| **Material pathway** | | | | |
| Material class | | | | <0.001 |
| Good physical living conditions | 1,915 (96.1%) | 5,584 (88.4%) | 553 (71.2%) | |
| Overcrowding with lower risk of poor housing conditions | 15 (0.8%) | 225 (3.6%) | 87 (11.2%) | |
| Poor housing conditions with lower risk of overcrowding | 62 (3.1%) | 510 (8.1%) | 137 (17.6%) | |
| **Household adversity pathway** | | | | |
| Domestic violence | | | | 0.003 |
| No | 1,642 (82.5%) | 5,053 (80.0%) | 588 (75.7%) | |
| Yes | 349 (17.5%) | 1,265 (20.0%) | 189 (24.3%) | |
| Substance misuse | | | | <0.001 |
| No | 1,855 (93.1%) | 5,706 (90.3%) | 673 (86.7%) | |
| Yes | 137 (6.9%) | 612 (9.7%) | 104 (13.3%) | |
| Maternal mental distress | 2.6 (2.5) | 3.0 (3.5) | 3.7 (4.2) | 0.075 |
| **Parent-child relations pathway** | | | | |
| Disciplinary practices | 13.3 (4.4) | 13.3 (4.8) | 12.1 (5.5) | <0.001 |
| Parent-child relationship | 50.1 (6.1) | 49.5 (6.7) | 47.7 (7.4) | <0.001 |
| Parenting activities | 33.7 (6.8) | 31.7 (7.3) | 29.8 (7.5) | <0.001 |

[1] n (%); Mean (SD)

[2] chi-squared test with Rao & Scott's second-order correction; Wilcoxon rank-sum test for complex survey samples

**Table S7. Survey-weighted mediator distribution of exposure status 17y complete case sample (Unweighted n=5612)**

| Characteristic | High, N = 1,182[1] | Middle, N = 3,320[1] | Low, N = 375[1] | p-value[2] |
|---|---|---|---|---|
| **Neighbourhood pathway** | | | | |
| Neighbourhood class | | | | <0.001 |
| Good conditions | 1,043 (88.3%) | 2,689 (81.0%) | 264 (70.5%) | |
| Poor conditions | 139 (11.7%) | 630 (19.0%) | 111 (29.5%) | |
| **Services and social support pathway** | | | | |
| Attended antenatal classes | | | | <0.001 |
| No | 503 (42.5%) | 1,980 (59.6%) | 314 (83.7%) | |
| Yes | 679 (57.5%) | 1,340 (40.4%) | 61 (16.3%) | |
| Attended 6 months centre-based childcare by 32 months | | | | <0.001 |
| No | 725 (61.4%) | 2,701 (81.4%) | 348 (92.8%) | |
| Yes | 456 (38.6%) | 619 (18.6%) | 27 (7.2%) | |
| Low social support | | | | <0.001 |
| No | 1,095 (92.7%) | 3,196 (96.3%) | 355 (94.6%) | |
| Yes | 86 (7.3%) | 124 (3.7%) | 20 (5.4%) | |
| **Material pathway** | | | | |
| Material class | | | | <0.001 |
| Good physical living conditions | 1,134 (96.0%) | 2,987 (90.0%) | 265 (70.6%) | |
| Overcrowding with lower risk of poor housing conditions | 7 (0.6%) | 87 (2.6%) | 50 (13.2%) | |
| Poor housing conditions with lower risk of overcrowding | 41 (3.5%) | 246 (7.4%) | 61 (16.2%) | |
| **Household adversity pathway** | | | | |
| Domestic violence | | | | 0.047 |
| No | 985 (83.4%) | 2,633 (79.3%) | 300 (80.1%) | |
| Yes | 196 (16.6%) | 686 (20.7%) | 75 (19.9%) | |
| Substance misuse | | | | 0.100 |
| No | 1,098 (92.9%) | 3,009 (90.7%) | 334 (89.1%) | |
| Yes | 84 (7.1%) | 310 (9.3%) | 41 (10.9%) | |
| Maternal mental distress | 2.6 (2.4) | 2.9 (3.2) | 3.1 (3.7) | 0.800 |
| **Parent-child relational pathway** | | | | |
| Disciplinary practices | 13.4 (4.2) | 13.2 (4.9) | 12.4 (5.7) | 0.086 |
| Parent-child relationship | 50.2 (6.1) | 49.6 (6.5) | 47.6 (7.3) | <0.001 |
| Parenting activities | 33.7 (6.7) | 32.0 (7.3) | 29.7 (7.5) | <0.001 |
| [1] n (%); Mean (SD) | | | | |
| [2] chi-squared test with Rao & Scott's second-order correction; Wilcoxon rank-sum test for complex survey samples | | | | |

**Table S8. Associations between Exposure-Outcome and Mediator-Outcome, separately adjusted for each exposure and mediator variable. Survey-weighted complete case analysis.**

| Predictor | Parent-reported mental health 5y (n=8477) | Parent-reported mental health 17y (n=5612) |
|---|---|---|
| **Childhood socioeconomic circumstances [Ref: High]** | | |
| Middle | 1.14 [0.89;1.40] *** | 1.28 [0.83;1.73] *** |
| Low | 2.48 [1.99;2.97] *** | 2.27 [1.20;3.34] *** |
| **Neighbourhood pathway** | | |
| Neighbourhood class [Ref: Good conditions] | | |
| Poor conditions | 1.10 [0.79;1.41] *** | 2.07 [1.22;2.93] *** |
| **Early support pathway** | | |
| Attended antenatal classes [Ref: No] | -0.15 [-0.37;0.07] | -0.01 [-0.56;0.55] |
| Attended centre-based childcare [Ref: No] | -0.05 [-0.33;0.22] | -0.19 [-0.67;0.29] |
| Low social support [Ref: No] | 0.81 [0.22;1.39] ** | 0.93 [0.14;1.72] * |
| **Home environment pathway** | | |
| Material class [Ref: Good physical conditions] | | |
| Overcrowding with lower risk of poor housing conditions | 1.19 [0.53;1.86] *** | 1.23 [-0.68;3.13] |
| Poor housing conditions with lower risk of overcrowding | 1.23 [0.73;1.74] *** | 1.02 [0.01;2.02] * |
| **Household adversity pathway** | | |
| Domestic violence [Ref: No] | 0.41 [0.14;0.68] ** | 1.07 [0.42;1.72] ** |
| Substance misuse [Ref: No] | 0.29 [-0.07;0.64] | -0.22 [-0.81;0.36] |
| Maternal mental distress | 0.3 [0.26;0.34] *** | 0.28 [0.21;0.35] *** |
| **Parent-child relational pathway** | | |
| Disciplinary practices | 0.16 [0.14;0.18] *** | 0.10 [0.05;0.15] *** |
| Parent-child relationship | -0.24 [-0.26;-0.22] *** | -0.16 [-0.19;-0.13] *** |
| Parenting activities | -0.07 [-0.09;-0.06] *** | -0.04 [-0.07;-0.00] * |

Note: Each exposure and mediating variable was estimated separately with the following adjustment sets:
*Exposure: sex, ethnicity, maternal age, maternal mental health history, maternal childhood adversity (care experience)*
*Mediators: additionally socioeconomic circumstances, all mediators in an earlier pathway, children in household, language spoken at home, gestational age, early developmental delays, long-term-limiting illness, parental high-frequency alcohol use*
*Mediator-Outcome associations are not adjusted for mediators in the same pathway.*

**Table S9. Sensitivity analyses - associations between Exposure-Outcome and Mediator-Outcome (MI), separately adjusted for each exposure and mediator variable. Multiply imputed data (m=10) with pooled estimates**

| Predictor | Self-reported mental health 17y (n=15513) | Parent-reported mental health 17y screen (Total difficulties ≥17) (n=15513) |
|---|---|---|
| **Childhood socioeconomic circumstances** | | |
| Maternal education [Ref: High] | | |
| Middle | 0.81 [0.55;1.07] *** | 1.75 [1.35;2.27] *** |
| Low | 1.39 [1.03;1.75] *** | 2.21 [1.65;2.97] *** |
| **Neighbourhood pathway** | | |
| Neighbourhood class [Ref: Good conditions] | | |
| Poor conditions | 0.74 [0.51;0.97] *** | 1.54 [1.32;1.8] *** |
| **Services and social support pathway** | | |
| Attended antenatal classes [Ref: No] | -0.48 [-0.79;-0.17] ** | 0.99 [0.83;1.18] |
| Attended centre-based childcare [Ref: No] | -0.26 [-0.58;0.06] | 1.02 [0.81;1.28] |
| Low social support [Ref: No] | 0.1 [-0.45;0.66] | 1.67 [1.22;2.29] ** |
| **Home environment pathway** | | |
| Material class [Ref: Good physical living conditions] | | |
| Overcrowding with lower risk of poor housing conditions | 0.45 [0;0.91] | 1.69 [1.32;2.18] *** |
| Poor housing conditions with lower risk of overcrowding | 0.61 [0.1;1.12] * | 1.24 [0.99;1.54] |
| **Household adversity pathway** | | |
| Domestic violence [Ref: No] | 0.5 [0.17;0.83] ** | 1.32 [1.09;1.59] ** |
| Substance misuse [Ref: No] | 0.4 [-0.03;0.84] | 1.04 [0.79;1.37] |
| Maternal mental distress | 0.08 [0.05;0.12] *** | 1.05 [1.03;1.07] *** |
| **Parent-child relational pathway** | | |
| Disciplinary practices | 0.03 [0;0.07] * | 1.03 [1.01;1.05] ** |
| Parent-child relationship | -0.07 [-0.09;-0.05] *** | 0.96 [0.94;0.97] *** |
| Parenting activities | 0.02 [0;0.03] * | 0.99 [0.98;1] * |

Note: Each exposure and mediating variable was estimated separately with the following adjustment sets:
*Exposure: sex, ethnicity, maternal age, maternal mental health history, maternal childhood adversity (care experience)*
*Mediators: additionally socioeconomic circumstances, all mediators in an earlier pathway, children in household, language spoken at home, gestational age, early developmental delays, long-term-limiting illness, parental high-frequency alcohol use*
*Mediator-Outcome associations are not adjusted for mediators in the same pathway.*

**Table S10. Sensitivity analyses using 17y self-reported mental health and binary mental health screen. Interventional Disparity Measures, Monte Carlo simulations with multiply imputed data (n=3,102,600 Monte Carlo observations)**

| | Self-reported mental health 17y | | Parent-reported mental health 17y screen (Total difficulties ≥17) | |
|---|---|---|---|---|
| **IDM** | **Score difference (Low vs High SEC)** | **% Total association** | **Risk difference (Low vs High SEC)** | **% Total association** |
| Total association (AdjTA) | 1.49 [1.27;2.03] | 100% | 0.091 [0.072;0.11] | 100% |
| Indirect effect (IE) | -0.63 [-1.07;-0.46] | -42% [-73;-20] | -0.042 [-0.062;-0.034] | -47% [-69;-35] |
| Residual association (RA) | 0.86 [0.4;1.37] | 58% [27;80] | 0.048 [0.021;0.065] | 53% [31;65] |
| **Indirect effect via five early childhood pathways** | | | | |
| Neighbourhood | -0.11 [-0.21;-0.02] | -8% [-15;1] | -0.001 [-0.005;0.006] | -1% [-5;7] |
| Services/social support | -0.32 [-0.68;-0.19] | -22% [-47;-7] | -0.016 [-0.036;-0.011] | -17% [-39;-12] |
| Material home environment | -0.06 [-0.16;0.08] | -4% [-12;8] | -0.008 [-0.015;-0.002] | -9% [-17;-1] |
| Household adversity | -0.01 [-0.06;0.06] | -1% [-5;5] | -0.001 [-0.003;0.003] | -1% [-4;3] |
| Parent-child relational | -0.12 [-0.37;0] | -8% [-26;3] | -0.017 [-0.026;-0.007] | -19% [-28;-8] |
| Mediated dependence | 0 [-0.003;0.002] | 0% [-2;3] | 0 [-0.03;0.04] | 0% [-3;3] |
